# Zirconia-based versus metal-framework veneered complete-arch implant-supported fixed dental prostheses: a systematic review of comparative clinical studies with integrated cost-effectiveness analysis

**DOI:** 10.64898/2026.09.02.26362012

**Authors:** Edward Coote, Nitesh Patel, Rajesh Vijayanarayanan

**Affiliations:** 21D Clinical Limited, Warrington, United Kingdom

**Keywords:** implant-supported fixed complete dental prosthesis, monolithic zirconia, titanium framework, PMMA, systematic review, cost-effectiveness analysis, All-on-4

## Abstract

**Objectives:** To systematically compare the outcomes of zirconia-based versus metal-framework veneered implant-supported fixed complete dental prostheses (ISFCDPs). We integrate cost-effectiveness analysis comparing monolithic zirconia versus titanium-PMMA ISFCDPs to model the 20-year costs and benefits associated with each arm.

**Data sources:** PubMed/MEDLINE, Cochrane CENTRAL, Scopus, ClinicalTrials.gov and WHO ICTRP were searched on March 6 2026 with no date restrictions applied.

**Study selection:** Two-arm comparative studies reporting clinical outcomes for both zirconia-based and metal-framework veneered ISFCDPs with a minimum of one-year follow-up after prosthesis delivery.

**Data extraction and synthesis:** Two independent reviewers extracted data and assessed the risk of bias using the Newcastle–Ottawa Scale. Narrative synthesis followed SWiM guidelines. A tiered material classification system was employed to handle framework alloy heterogeneity. Meta-analysis was not possible due to heterogeneity in outcomes reported and methodological differences. A Markov cohort model estimated cost-effectiveness from a patient perspective over a 20-year horizon. Deterministic and probabilistic sensitivity analysis (PSA) tested results robustness.

**Results:** Eight retrospective cohort studies (396 patients; 551 prostheses) were included in final analysis. All were classified as Tier 1 due to inconsistent alloy reporting. Prosthesis and implant survival rates were high across both arms but with no statistical difference for any primary outcome. Cost-effectiveness analysis estimated total discounted per-patient costs of £23,295 for titanium-PMMA and £29,170 for monolithic zirconia over 20 years. This results in an ICER of £315,417 per QALY gained. PSA confirmed zirconia as cost-effective in only 0.50% of 10,000 iterations at the £20,000/QALY threshold. Net monetary benefit was consistently negative for zirconia.

**Conclusions:** Both types of ISFCDPs provided high prosthesis and implant survival with no statistically significant difference in primary clinical outcomes, though all evidence was graded at very low certainty. We estimated a per-patient premium of £5,875 for monolithic zirconia over a 20-year horizon with marginal QALY gains (0.019). Adequately powered RCTs with standardised material reporting are needed to strengthen our clinical and economic conclusions.

**Systematic review registration:** OSF: *10.17605/OSFJO/CF796*

## 1. INTRODUCTION

Current global assessments of edentulism (defined as total tooth loss) estimate the global average prevalence at 7% among people aged 20 years and over, with this steeply rising to a much higher prevalence of 23% for people aged 60 years or older. A further 1 billion are estimated to suffer from severe periodontal diseases, which places them at immediate risk of complete tooth loss [1]. A 2024 systematic review across country income groups (low-, middle- and high-income) reported the considerable impact that edentulism has on health and socioeconomic outcomes, such as poorer nutrition, quality of life and general and mental health [2]. As a result, it is expected that those who can access (subject to affordability and availability of clinics and suitable dental clinicians) treatment will seek it out. Complete-arch implant-supported fixed complete dental prostheses (ISFCDPs) are established as the standard of care for edentulous full-jaw rehabilitation, typically using the All-on-Four concept, since the demonstration of osseointegration by Brånemark and colleagues and subsequent longitudinal data to demonstrate their longevity and benefits to patients [3, 4, 5].

Advances in computer-aided design and computer-aided manufacturing (CAD/CAM) alongside high survival and success rates of dental implants has resulted in two dominant ISFCDP concepts [6]: two-component metal framework (using different alloys such as titanium, cobalt-chromium and nickel-chromium) with an acrylic/PMMA structure forming the teeth and gingiva, and an implant-supported zirconia-based prosthesis structure [7]. There are advantages and disadvantages to both methods which can cause confusion to both patient and clinician when determining which is best. Metal-acrylic combinations have been long standing due to their high prosthetic survival rates, affordability, and biocompatibility [8]. However, recent advances in dental manufacturing technology and the increased demand for aesthetic solutions has resulted in an increase in demand for all-ceramic reconstructions such as monolithic zirconia ISFCDPs [9]. The 2018 International Team for Implantology (ITI) stated that “implant-supported monolithic zirconia prostheses may be a future option with more supporting evidence”, highlighting the need for more randomised controlled trials (RCTs) and better evidence synthesis [10].

Existing reviews have focused on single material categories at a time, limiting head-to-head comparability, been limited to scoping or narrative reviews, or focused on single-arm studies of prosthetics [7, 8, 11]. There is limited observational evidence that is characterised by heterogeneity in prosthetic materials with studies often being ambiguous and general (stating metal-acrylic). Due to the differences in the performances of certain material combinations, there is a need for strong evidence for specifics. For example, there is only one RCT protocol registered comparing zirconia frameworks with ceramic veneering versus titanium frameworks with acrylic resin veneering. Its primary outcome is peri-implant submucosal microbiota diversity [12]. Currently, there is limited cost-effectiveness analysis (CEA) comparing full-arch prosthetic approaches. In the context of constrained healthcare budgets, different dental payment systems, and differences in manufacturing costs it is important to provide a tool to assess the two prosthetic approaches which can inform clinical decisions.

In this review, we aim to systematically compare the clinical outcomes of zirconia-based ISFCDPs versus metal-framework veneered ISFCDP. We developed a three-tiered material classification system for the comparator arm to manage the heterogeneity in framework reporting. This ranges from any metal-framework with any veneering material (broadest evidence capture), to CAD/CAM titanium framework with PMMA veneering (the specific contemporary comparison of interest). Subgroup analysis by zirconia design type (monolithic versus veneered) was utilised for the intervention arm. We compare secondary outcomes such as peri-implant health parameters and patient-reported outcomes (PROMs), and determine if differences in outcomes exist between variables such as jaw location (maxilla vs mandible), the framework alloy used, or by loading protocol (immediate vs delayed). Using this analysis, we will integrate cost data with the benefits of the two ISFCDPs to produce a Markov cost-effectiveness model to inform patient and clinical decisions.

## 2. METHODS

### 2.1 Protocol and registration

This systematic review was conducted in line with the Preferred Reporting Items for Systematic Review and Meta-Analysis (PRISMA) 2020 guidelines, with results following the Synthesis Without Meta-analysis (SWiM), and the economic evaluation following the Consolidated Health Economic Evaluation Reporting Standards (CHEERS) 2022 reporting checklist [13, 14, 15]. The protocol was registered on OSF (10.17605/OSFJO/CF796). Completed checklists are provided in the Supplementary Material (Table S1–S3).

### 2.2 Eligibility criteria

#### 2.2.1 Population

Adult patients (*≥*18 years) with one or both edentulous arches rehabilitated with complete-arch ISFCDPs on four or more implants (All-on-4, All-on-6).

#### 2.2.2 Intervention

Monolithic, predominately monolithic zirconia complete-arch ISFCDPs, or zirconia-framework with ceramic veneering, with zirconia providing the primary structural framework.

#### 2.2.3 Comparator

Metal-framework complete-arch ISFCDPs with polymer (PMMA, acrylic resin, composite) prosthetic teeth/gingiva. To include more studies and to avoid definition overlap, a tiered material classification was applied (see Section 2.8).

#### 2.2.4 Outcomes

**Primary outcomes:** prosthetic survival rate, bone resorption, biological and technical complications.

**Secondary outcomes:** implant survival, peri-implant clinical parameters (plaque index, bleeding on probing), PROMs, and cost data.

#### 2.2.5 Study designs

Eligible studies included: RCTs, controlled clinical trials, prospective and retrospective comparative cohort studies, and cross-sectional comparative studies. The key criteria was for studies to report both the intervention and comparator groups within the same study. A minimum of 1-year of follow up after prosthetic delivery was required.

#### 2.2.6 Exclusion criteria

Single-arm case studies, case reports, in vitro and animal studies, narrative and systematic reviews, papers not dealing with original clinical cases (such as reviews, conference abstracts, editorials). Reports based on questionnaires or interviews, where no attempt to clinically examine patients, were also excluded. A hand search of the reference list of relevant systematic and narrative reviews was performed to ensure the inclusion of all potentially relevant studies.

### 2.3 Information sources

The following Internet sources used to search for papers: National Library of Medicine (PubMed/MEDLINE), Embase, Cochrane CENTRAL and Scopus. Grey literature sources included ClinicalTrials.gov and WHO ICTRP. The last search was March 6 2026. As mentioned in Section 2.2.6, a hand search of the reference list of relevant reviews was also conducted. No date restriction was applied.

### 2.4 Search strategy

Our search strategy used three concept blocks using Boolean AND operators: (1) dental implants and complete-arch prosthetics terms, (2) zirconia and monolithic terms, (3) metal framework, titanium, PMMA and acrylic terms, adjusted to each database searches syntax. The comparator concept included all potential metal alloy terms to ensure an initial broad literature capture. We then later used the tiered material classification (see Section 2.8) during data extraction (rather than at the search stage) to filter down the results. The search strategy was validated against five known eligible studies identified during scoping [16, 17, 18, 19, 20]. This validation set includes studies that we expect our search to retrieve and does not confirm they will be included in the final included studies after full-text screening. Full database-specific search strings are provided in the Supplementary Material (Table S4).

### 2.5 Selection process

All results from the five databases were imported into Rayyan to deduplicate and then title/abstract screening was performed by two independent reviewers (E.C. and N.P.). Disagreement between the reviewers were resolved through discussion with R.V. being used as a moderator. Cohen’s *κ* between the two independent reviewers was 0.23, which was mainly due to disagreement over which outcomes to include (decision to not use studies that only measure stress distributions or an implant not prosthesis focus, the third review was not required due to discussion between E.C. and N.P.). Due to the expectation of a low number of final studies, a liberal inclusion approach at title/abstract screening was applied. Papers marked as unclear advanced to full-text assessment due to potential vagueness in their title and abstract. At the full-text stage, both reviewers independently assessed each study against the eligibility criteria, with a third reviewer not required.

### 2.6 Data collection process

Data items from individual studies included: study characteristics, population demographics, implant characteristics, prosthesis material specification (framework alloy, fabrication method, veneering material, fabrication method), along with all primary and secondary outcomes. Each study was assigned a material tier classification at extraction (See Section 2.8). If a study reported ‘metal framework’ without specifying the alloy, corresponding authors were contacted for clarification.

### 2.7 Outcomes and prioritisation

Prosthetic survival was defined as the prosthesis remaining in situ without modifications (from the first and only implant surgery) [21]. Technical complications were categorised as: prosthetic framework or teeth/gingiva fracture, screw loosening or fracture, prosthesis remake and subsequent rounds of surgery to re-fit. Biological complications included: peri-implantitis, marginal bone loss, plaque accumulation and soft tissue complications.

### 2.8 Tiered material classification

Due to the serious heterogeneity in framework alloy reporting across the literature, we developed a three-tiered classification system to manage this heterogeneity in reporting. Many studies in the literature currently report using ‘metal-acrylic’ or reviews group all metal frameworks (titanium, cobalt-chromium) together, despite key biocompatibility and corrosion differences [8, 22]. This was an attempt to reduce the introduction of unmeasured heterogeneity that would distort the true differences against the intervention. Tier 1 was designed as the broadest evidence capture to maximise the potential relevant studies assessed due to the reporting problem specified. Tier 2 and Tier 3 narrows our scope to more clinical specifics and will inform our CEA. Performing our review across using these Tiers allowed us to determine whether our conclusions are the same across all three tiers with results robust to the framework alloy specification problem. The tier system is only applicable to the comparator arm, while differing levels of the intervention arm (such as monolithic versus veneered zirconia) will be assessed in subgroups.

### 2.9 Risk of bias assessment

We used the Newcastle–Ottawa Scale (NOS) to assess non-randomised comparative cohort studies, and an adapted version for cross-sectional studies if needed [23]. The risk of bias was assessed independently using two reviewers (E.C. and N.P.). Outcomes are reported in Figure 2.

### 2.10 Data synthesis

#### 2.10.1 Narrative synthesis

All included studies were described narratively which followed the SWiM reporting guidelines [13]. We created an evidence gap matrix in Table 4 to map which outcomes were reported across which studies. This helped us to identify evidence gaps and data saturation.

#### 2.10.2 Quantitative synthesis

Meta-analysis was planned for scenarios where two or more studies reported the same outcome using comparable definitions, populations and follow-up durations [24]. For example, prosthesis survival at 5 years. For studies that were considered sufficiently homogeneous, random-effects meta-analysis using the DerSimonian and Laird method was used [25].

### 2.11 Certainty of evidence

We used the Grading of Recommendations, Assessment, Development and Evaluations (GRADE) framework to assess the certainty of the body of evidence for each outcome [27]. We assessed risk of bias, inconsistency, indirectness, imprecision and publication bias. These categories were then used to assess certainty of evidence as high, moderate, low, or very low (results presented in Table 6).

### 2.12 Cost-effectiveness analysis

#### 2.12.1 Model structure

We produced a decision-analytics model to estimate the long-term costs and health outcomes associated with monolithic zirconia versus titanium-PMMA (Ti-PMMA) ISFCDPs for complete-arch rehabilitation. We developed an initial decision tree which reflects a patient/clinician choice for treatment selection and first years events, followed by a Markov cohort model simulating a horizon of 20 years (cycle length of one year) and 1,000 individuals to capture the prosthetic life span. Each arm of the decision tree will capture the initial fabrication cost. Patients then exit the decision tree and enter the Markov model with a functioning prosthesis. The Markov model will be composed of five mutually exclusive health states: (1) functioning and intact prosthetics that receives routine annual maintenance once a year, (2) a prosthetic with minor complications requiring chairside or short laboratory repair such as veneering chipping or screw loosening, (3) major complications that require complete remake of prosthesis due to serious breakage such as framework fracture or irreparable delamination, (4) implant failure of one or more supporting implants that need clinical management, (5) complete prosthesis failure leading to abandonment of ISFCDPs with transition to removable dentures or edentulism. At each cycle, patients are able to transition between the health states using probabilities that have been derived from our systematic review. Patients that experience minor complications returned to the functioning prosthetic state after repair. Patients that experienced major complications returned to the functioning prosthetic state after full remake, which includes a full fabrication cost again. Prosthesis failure was an absorbing state. Half-cycle corrections were applied to reoccurring Markov costs and QALYs from cycle 1 onwards. Transition probabilities for the two arms were expected to differ given the distinct profiles and characteristics of each ISFCDP. Tier 2 and Tier 3 data from the systematic review was used when sufficient evidence was provided. Tier 1 pooled estimates were used in our sensitivity analysis. The model is from a per-patient perspective in which all patients received both arch full-jaw rehabilitation.

#### 2.12.2 Perspective and discounting

We took the patient perspective for costs as full-jaw rehabilitation is not covered by the NHS. This includes travel costs, time off work for appointments and surgery and out-of-pocket repair costs. The commercial cost of the full treatment include implant placement surgery and prosthetic placement surgery. We assumed immediate loading of final long-term prosthetics on All-on-4 prosthetics. As a form of sensitivity analysis we varied a copayment module which reduced the direct healthcare costs for patients to see the effect on the final incremental cost effectiveness ratio (ICER). Costs and outcomes were discounted at 3.5% per annum following National Institute of Care and Excellence guidance [28]. Scenario analyses varied the discount rate at 0%, 2% and 5%.

#### 2.12.3 Clinical parameters

Where our systematic review failed to provide sufficient data for transition probabilities, supplementary literature was used. Otherwise, clinical transition probabilities were primarily derived from our review. Tier 1 data was prioritised, and pooled Tier 2 and Tier 3 data used in our sensitivity analysis. All parameter values, ranges, distributions and sources can be found in Table 7.

#### 2.12.4 Cost inputs

Initial fabrication for both intervention arms were derived from surveying multiple realworld industry providers (Supplementary Material Tables S5). Maintenance and repair costs were added separately. Travel costs were estimated using HMRC cars and vans mileage rates and the average distance from a specialist full-jaw clinic [29]. The ONS Annual Survey of Hours and Earnings median hourly wage was multiplied by the estimated hours per appointment for initial delivery, minor repair and major prosthetic replacement and annual maintenance [30]. Full cost (at 2025£ prices) are found in Table 7. Costs are varied greatly in scenario sensitivity analysis.

#### 2.12.5 Health outcomes

We use quality-adjusted life years (QALYs) in our model, derived from age-sex matched UK general population norms as there are currently no published EQ-5D utility values specific to ISFCDPs [31]. Therefore, our baseline utility for the functioning prosthesis (state 1) was estimated using the Ara and Brazier [32] regression equation. The baseline utility value was determined to be 0.83 given a starting age 60 years and a mixed-sex cohort. We then applied a disutility to states that required one. Minor complications (state 2) had a disutility of *−*0.05, a disutility of *−*0.15 was applied for major complications (state 3), implant failure (state 4) had a disutility value of *−*0.20. The values have been justified in Supplementary Table S6. Complete prosthetic failure (state 5) had a disutility multiplier of 0.10 applied to the age-adjusted baseline so that it was always below the population norm. This is due to the absence of quality-of-life data for edentulous patients with conventional dentures [33, 34, 35, 36, 37]. Office for National Statistics life tables for England were used for all-cause mortality rates for the absorbing death state [38]. Full utility values and their detailed justification can be found in the Supplementary Material Table S6.

#### 2.12.6 Sensitivity and uncertainty analyses

All clinical parameters, cost inputs and health outcomes were varied using one-way deterministic sensitivity analysis and Monte Carlo simulations. Tornado diagrams were produced by varying the high and low value of each parameter. We ran 10,000 Monte Carlo simulations to build a cost-effectiveness plane and acceptability curve. Our scenario analyses included: varying the discount rate and testing copayment scenarios.

## 3. RESULTS

**Part 1: Systematic Review**

### 3.1 Study selection

Our initial database search yielded 3,539 total results, of which PubMed/MEDLINE produced 1,538, 1,781 from Scopus, 1 in Cochrane reviews, 144 in Cochrane trials, 30 from clinicaltrials.gov and WHO ICTRP produced 45. After duplicates were removed 2,053 articles remained. A total of 2,043 articles were excluded after title/abstract screening. This meant that 10 studies were left for full-text review after agreement between EC and NP. One study was excluded as it was a trial protocol article with the final results not yet published on clinicaltrial.gov [39]. The final selection was 8 studies [16, 17, 19, 20, 40, 41, 42, 43]. The PRISMA flow diagram summarising our search results is found in Figure 1. The majority of excluded studies were done so because they were single arm or not evaluating full-jaw prosthetics, only single tooth or section replacements.

**Figure 1:**
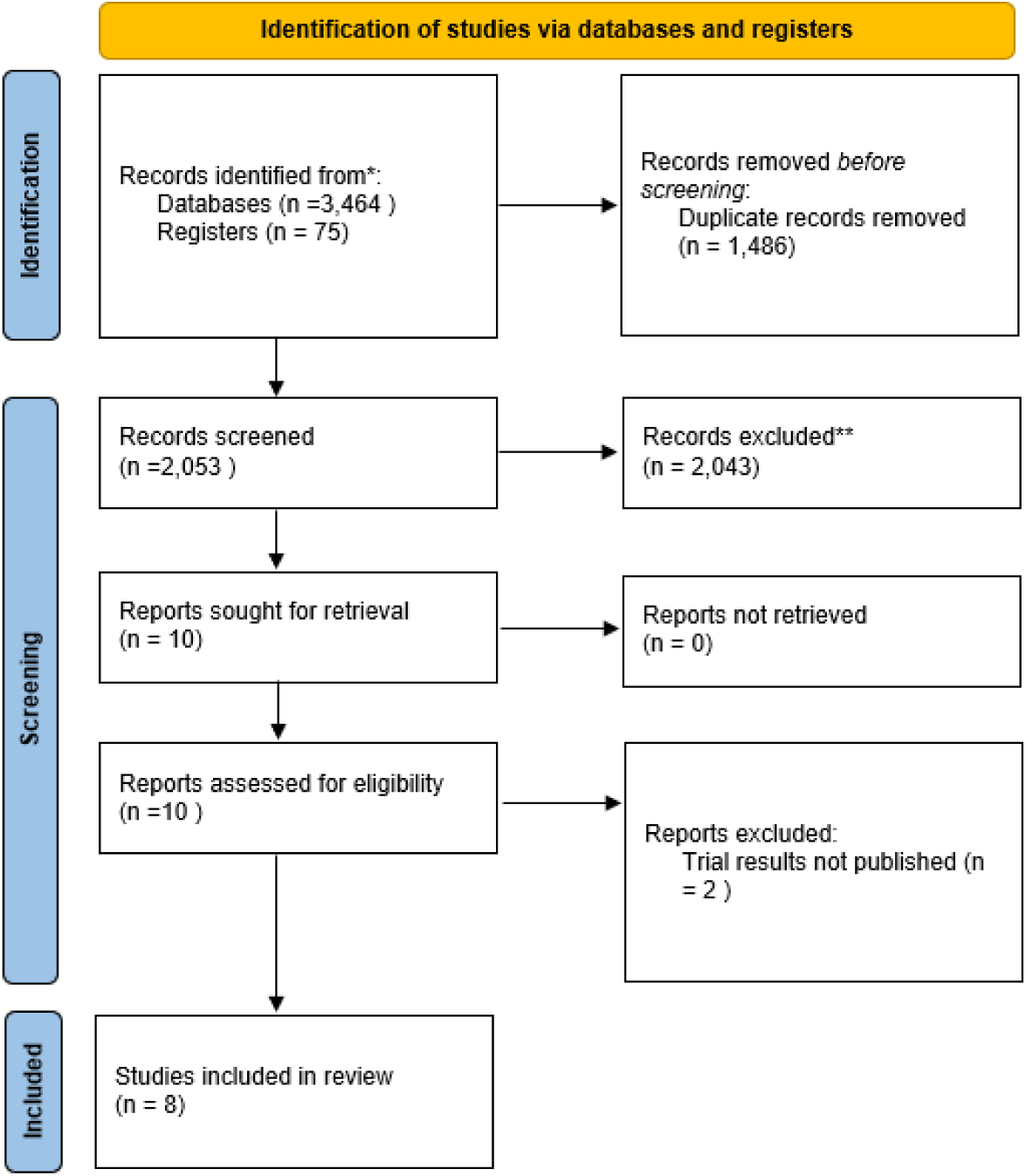
PRISMA 2020 flow diagram showing identification, screening, eligibility, and inclusion of studies. Adapted from Page et al. (2021) doi: 10.1136/bmj.n71.

**Figure 2:**
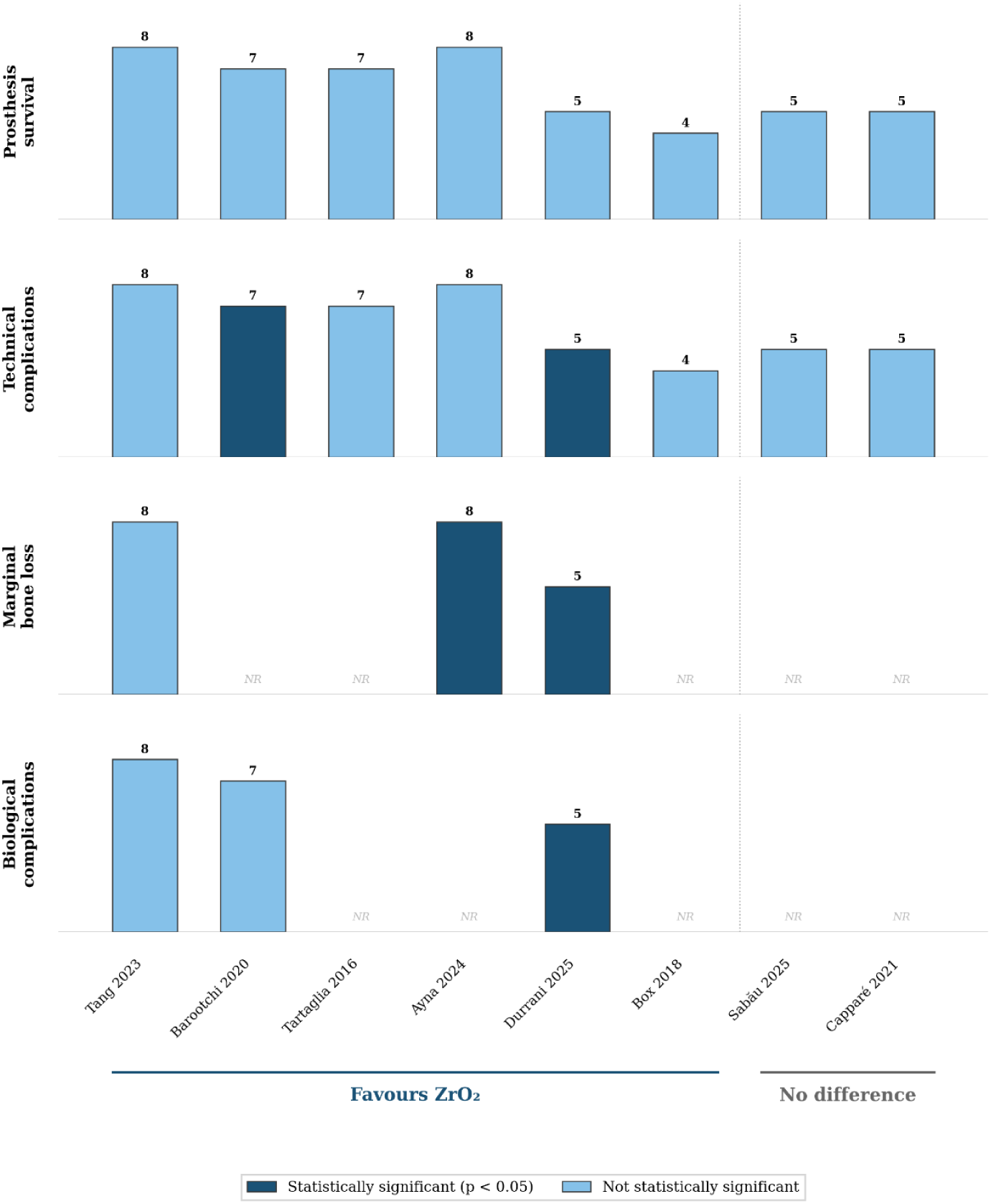
Harvest plot showing direction and magnitude of effects across studies for primary outcomes. Bar height proportional to NOS score (number above bar).

**Figure 3:**
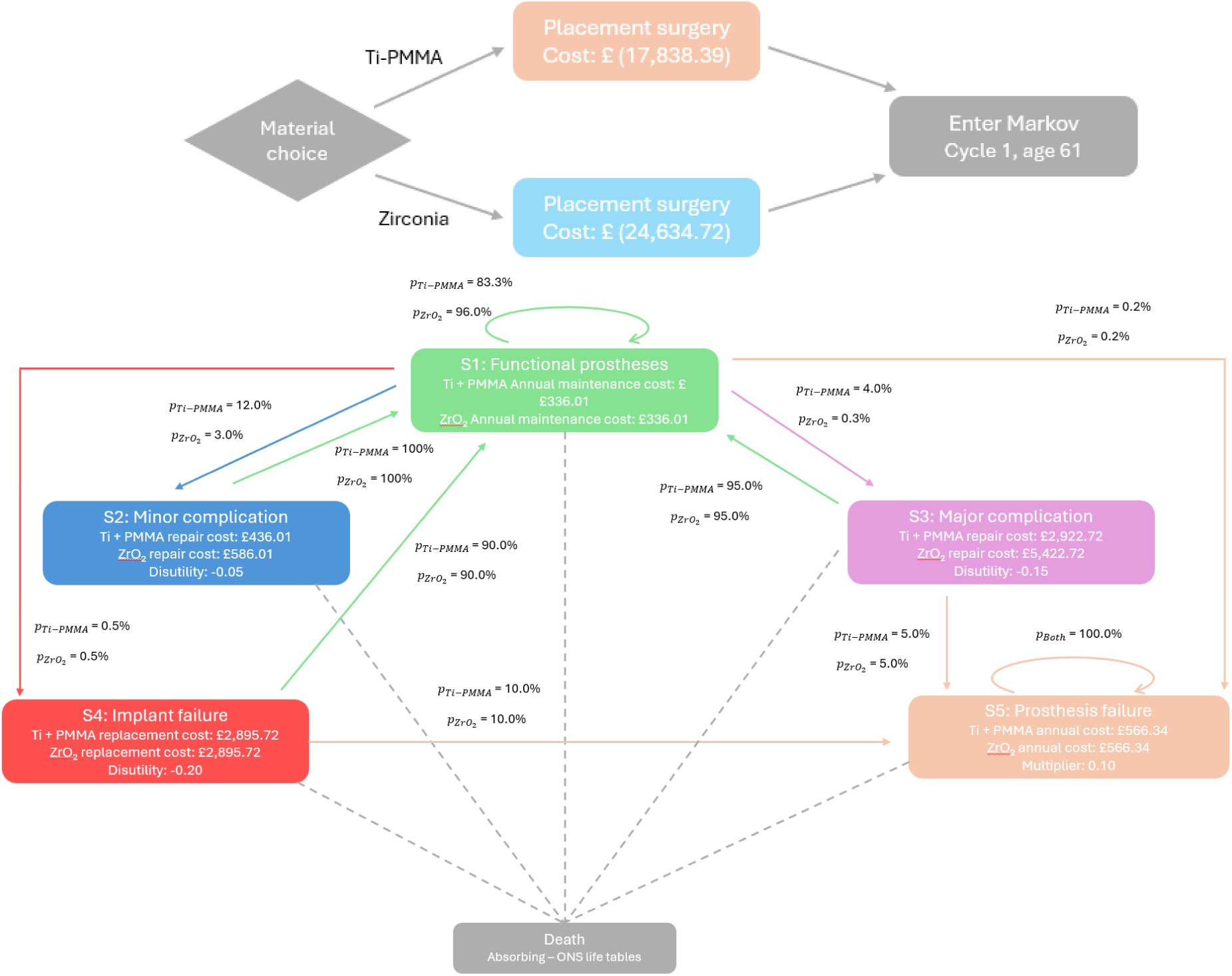
Markov model structure. Health states: functioning prosthesis, minor complication (repair), major complication (remake), implant loss, prosthesis failure/abandonment. Cycle length: 1 year. Arrows indicate permitted transitions between health states with their allocated annual transition probabilities.

### 3.2 Study characteristics

All eight selected studies were retrospective cohort studies from single centres (a mix of university, private practise and hospital), with only one being a retrospective analysis of multiple cases (full details in Table 3) [41]. Follow-up periods ranged from 2 months to 104 months, and prosthetic sample sizes ranged from 20 to 214. No meta-analysis for any outputs could be completed due to the vast heterogeneity across the studies, such as inconsistency in definitions of minor versus complications, the study originating from training/university institutions versus specialist practises, and the different unspecified metals used. There was heterogeneity in the zirconia arm (monolithic, predominantly monolithic, veneered), and in the metal-framework arm (different alloys used, unspecified alloys, fabrication processes). Only Tang, 2023 reported using a titanium framework, and we confirmed through email with Box, 2018 that a titanium framework was used [19, 43]. Other studies used a range of chromium based framework material, one using PMMA as the framework material, and only Barootchi, 2020 as unspecified metal. Five studies reported using acrylic resin or composite resin, although we were not able to confirm this as PMMA. Five studies used monolithic zirconia (Sabău, 2025; Durrani, 2025; Tang, 2023; Ayna, 2024; Box, 2018) two used veneered zirconia prostheses (Barootchi, 2020; Tartaglia, 2016) and one used only predominantly monolithic zirconia prostheses (Capparé, 2021). Two studies used a mixture of monolithic zirconia and partially monolithic (Sabău, 2025; Box, 2018), however separated results by prostheses sub-type. We decided to include Tartaglia et al (2015) as the comparator prosthesis has PMMA veneered with resin and a titanium bar framework, although described as a resin hybrid [20]. It still uses the framework-veneer structure comparable to metal-acrylic prosthesis. This study also provides evidence for the clinical performance of PMMA veneering, which is of utility given our PICO questions.

**Table 1:** Tiered material classification system for the metal-framework comparator arm.

| **Tier** | **Framework Alloy** | **Framework Fabrication** | **Veneering Material** | **Scope** |
| --- | --- | --- | --- | --- |
| 1 | Any metal<br>(including unspecified) | Any (including unspecified) | Any polymer<br>(PMMA, acrylic, composite, resin) | Primary analysis, broadest evidence capture |
| 2 | Titanium | Any (including unspecified) | PMMA or acrylic resin (confirmed) | Sensitivity analysis 1, alloy-specific |
| 3 | Titanium, CAD/CAM milled | CAD/CAM milled | PMMA, CAD/CAM milled | Sensitivity analysis 2; contemporary digital workflow match |

*Tier 1 serves as the primary analysis (broadest evidence capture). Tiers 2 and 3 are pre-specified sensitivity analyses progressively narrowing toward the specific contemporary clinical comparison. Studies where framework alloy could not be confirmed were included at Tier 1 only.*

**Table 2:** Risk of bias summary. Newcastle–Ottawa Scale star ratings for each included study across Selection, Comparability, and Outcome domains.

| Study | Selection |  |  |  | Comp. | Outcome |  |  |
| --- | --- | --- | --- | --- | --- | --- | --- | --- |
|  | Rep. | Non-exp. | Asc.exp. | Out.start | Comp.cohorts | Assess. | Follow-up | Adeq. |
| Sabău, 2025 | * | * | * | * | — | — | * | — |
| Durrani, 2025 | * | — | * | * | — | * | * | — |
| Tang, 2023 | * | * | * | * | ** | * | * | — |
| Ayna, 2024 | — | * | * | * | * | * | * | — |
| Barootchi, 2020 | * | * | * | * | ** | — | * | — |
| Tartaglia, 2015 | — | * | * | * | * | — | * | * |
| Box, 2018 | * | * | * | * | — | — | — | — |
| Capparé, 2021 | * | * | * | * | — | — | * | — |

**Table 3:**
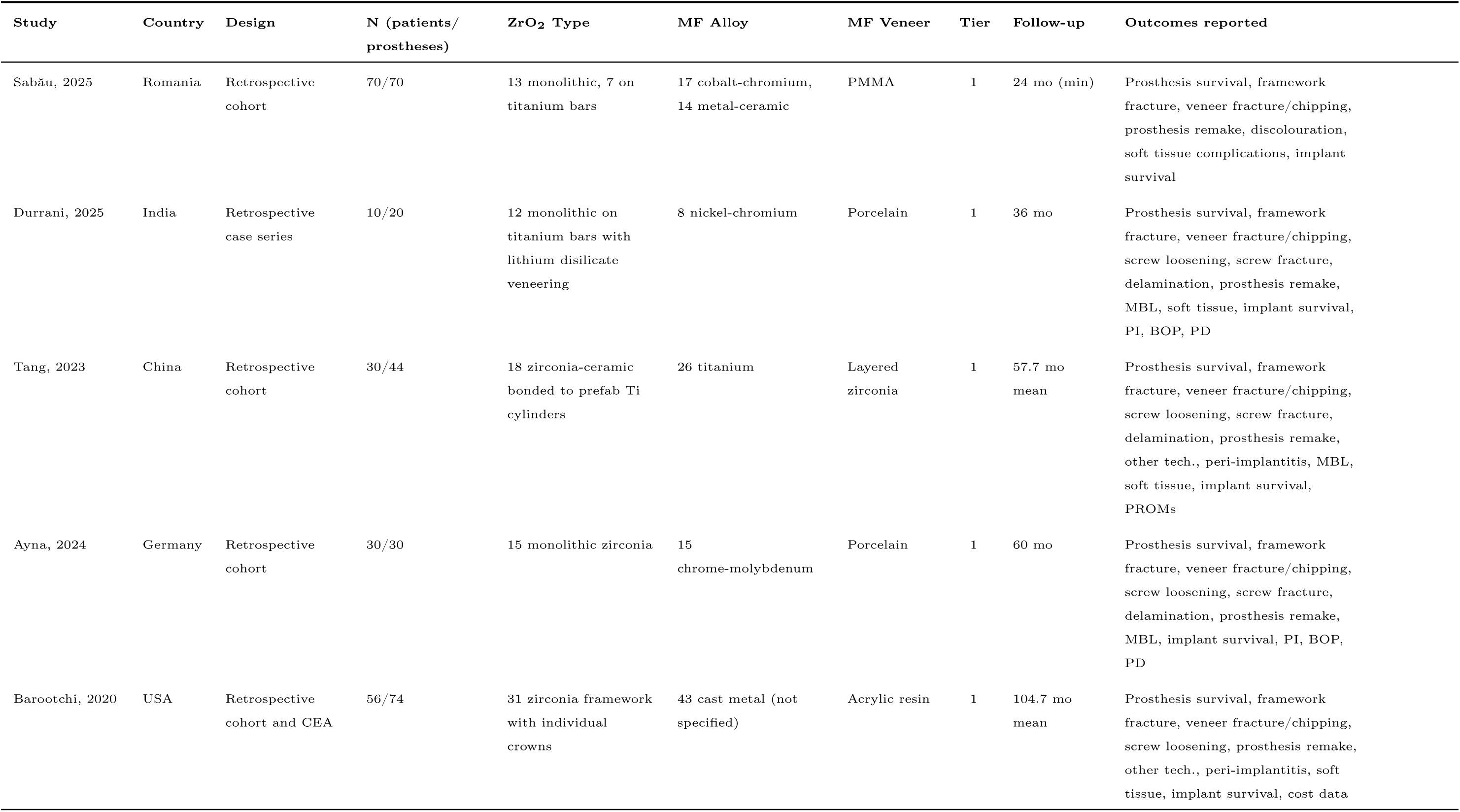
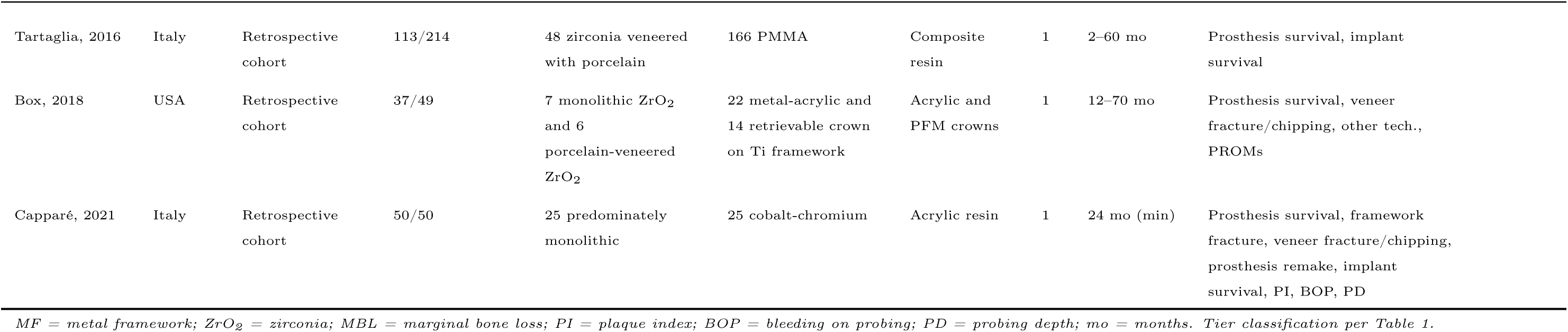
Characteristics of included studies.

| Study | Country | Design | N (patients/<br>prostheses) | ZrO<sub>2</sub> Type | MF Alloy | MF Veneer | Tier | Follow-up | Outcomes reported |
| --- | --- | --- | --- | --- | --- | --- | --- | --- | --- |
| Sabău, 2025 | Romania | Retrospective cohort | 70/70 | 13 monolithic, 7 on titanium bars | 17 cobalt-chromium, 14 metal-ceramic | PMMA | 1 | 24 mo (min) | Prosthesis survival, framework fracture, veneer fracture/chipping, prosthesis remake, discolouration, soft tissue complications, implant survival |
| Durrani, 2025 | India | Retrospective case series | 10/20 | 12 monolithic on titanium bars with lithium disilicate veneering | 8 nickel-chromium | Porcelain | 1 | 36 mo | Prosthesis survival, framework fracture, veneer fracture/chipping, screw loosening, screw fracture, delamination, prosthesis remake, MBL, soft tissue, implant survival, PI, BOP, PD |
| Tang, 2023 | China | Retrospective cohort | 30/44 | 18 zirconia-ceramic bonded to prefab Ti cylinders | 26 titanium | Layered zirconia | 1 | 57.7 mo mean | Prosthesis survival, framework fracture, veneer fracture/chipping, screw loosening, screw fracture, delamination, prosthesis remake, other tech., peri-implantitis, MBL, soft tissue, implant survival, PROMs |
| Ayna, 2024 | Germany | Retrospective cohort | 30/30 | 15 monolithic zirconia | 15 chrome-molybdenum | Porcelain | 1 | 60 mo | Prosthesis survival, framework fracture, veneer fracture/chipping, screw loosening, screw fracture, delamination, prosthesis remake, MBL, implant survival, PI, BOP, PD |
| Barootchi, 2020 | USA | Retrospective cohort and CEA | 56/74 | 31 zirconia framework with individual crowns | 43 cast metal (not specified) | Acrylic resin | 1 | 104.7 mo mean | Prosthesis survival, framework fracture, veneer fracture/chipping, screw loosening, prosthesis remake, other tech., peri-implantitis, soft tissue, implant survival, cost data |
| Tartaglia, 2016 | Italy | Retrospective cohort | 113/214 | 48 zirconia veneered with porcelain | 166 PMMA | Composite resin | 1 | 2–60 mo | Prosthesis survival, implant survival |
| Box, 2018 | USA | Retrospective cohort | 37/49 | 7 monolithic ZrO<sub>2</sub> and 6 porcelain-veneered ZrO<sub>2</sub> | 22 metal-acrylic and 14 retrievable crown on Ti framework | Acrylic and PFM crowns | 1 | 12–70 mo | Prosthesis survival, veneer fracture/chipping, other tech., PROMs |
| Capparé, 2021 | Italy | Retrospective cohort | 50/50 | 25 predominately monolithic | 25 cobalt-chromium | Acrylic resin | 1 | 24 mo (min) | Prosthesis survival, framework fracture, veneer fracture/chipping, prosthesis remake, implant survival, PI, BOP, PD |

*MF = metal framework; ZrO<sub>2</sub> = zirconia; MBL = marginal bone loss; PI = plaque index; BOP = bleeding on probing; PD = probing depth; mo = months. Tier classification per Table 1.*

**Table 4:**
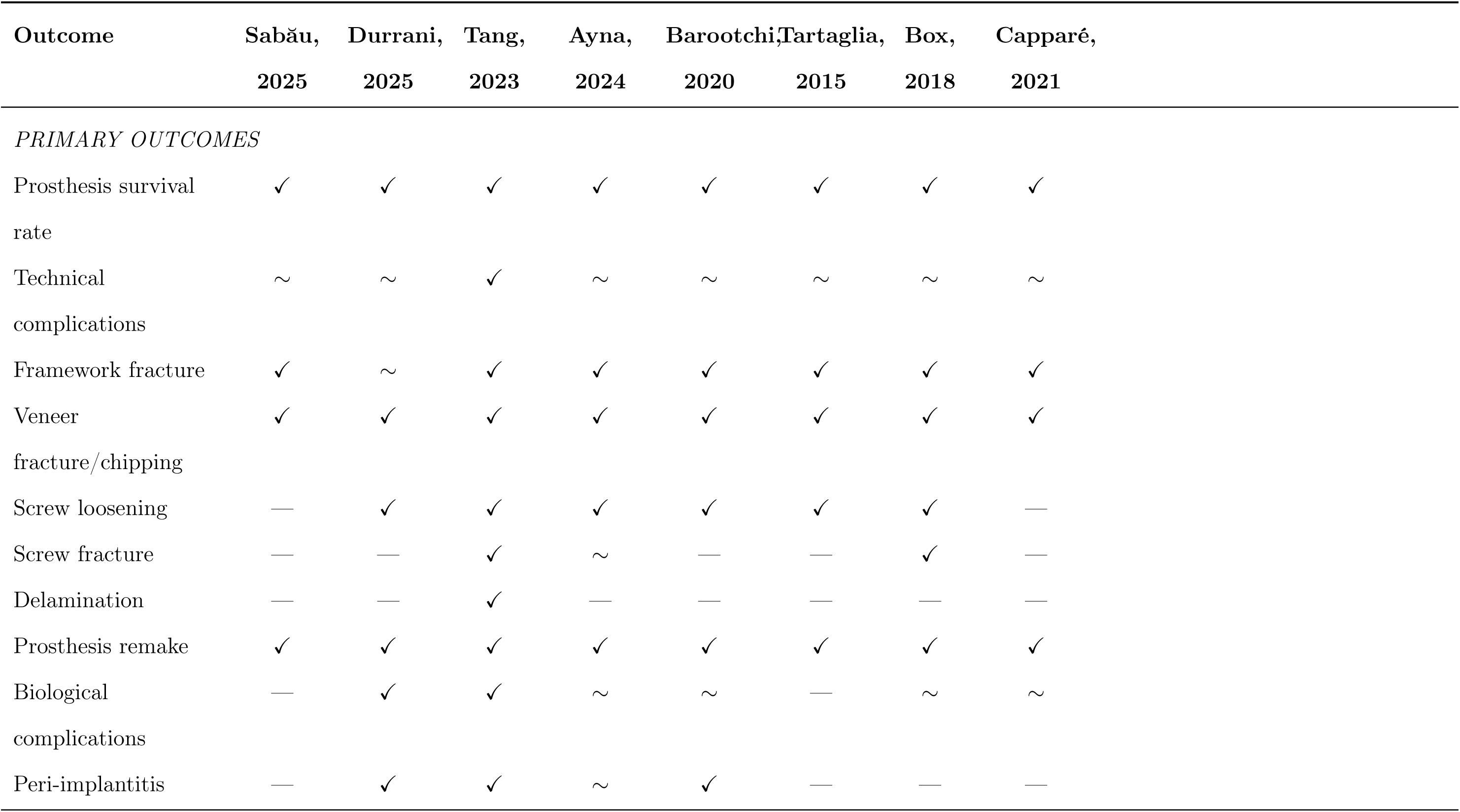
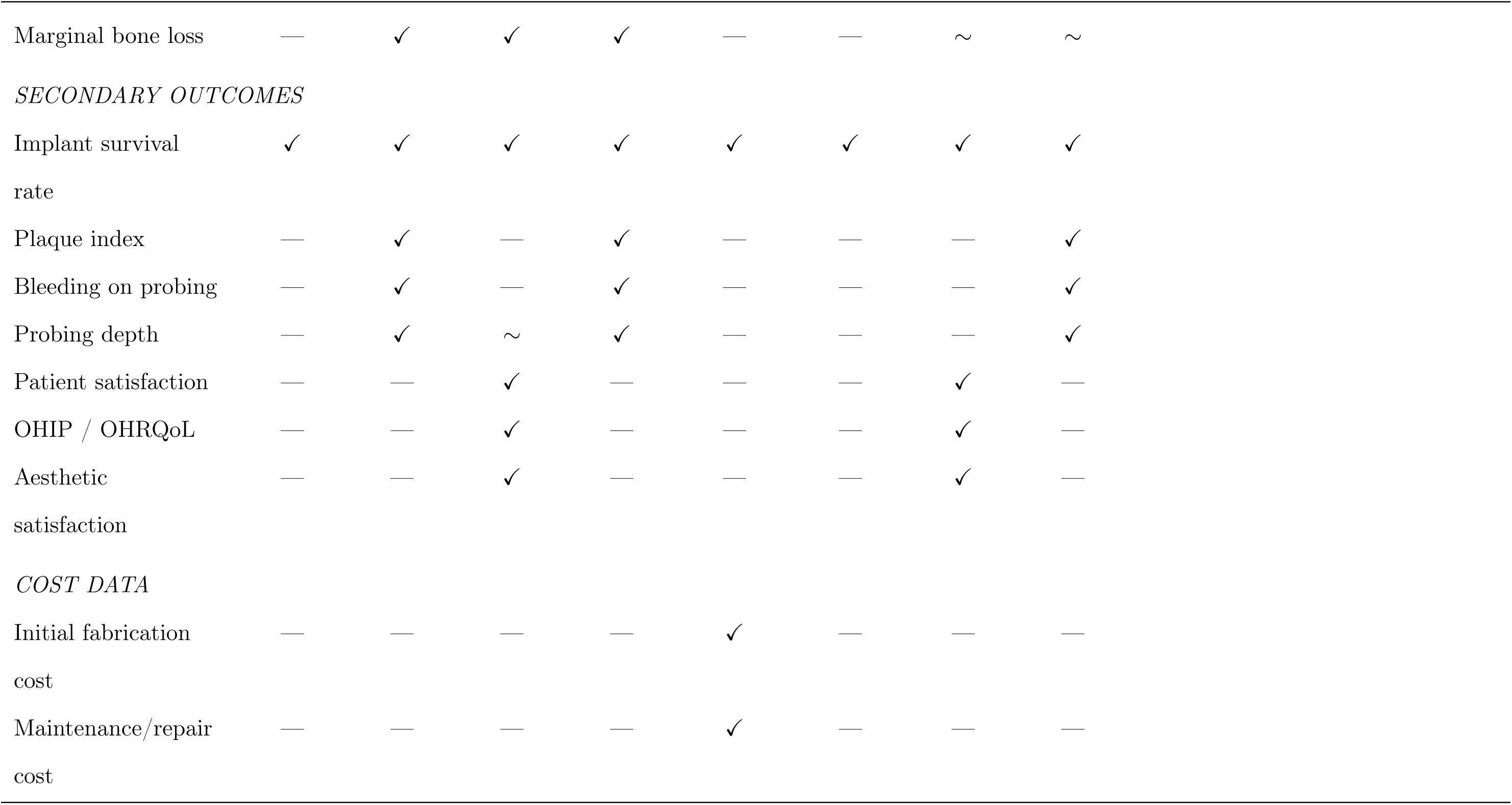
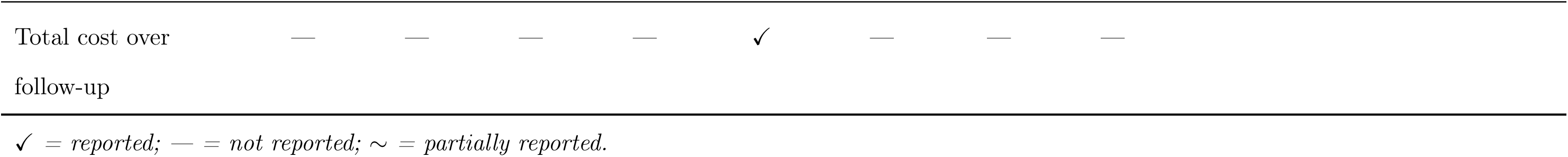
Evidence gap matrix: outcomes reported across included studies.

| Outcome | Sabău,<br>2025 | Durrani,<br>2025 | Tang,<br>2023 | Ayna,<br>2024 | Barootchi,<br>2020 | Tartaglia,<br>2015 | Box,<br>2018 | Capparé,<br>2021 |
| --- | --- | --- | --- | --- | --- | --- | --- | --- |
| *PRIMARY OUTCOMES* |  |  |  |  |  |  |  |  |
| Prosthesis survival rate | ✓ | ✓ | ✓ | ✓ | ✓ | ✓ | ✓ | ✓ |
| Technical complications | ~ | ~ | ✓ | ~ | ~ | ~ | ~ | ~ |
| Framework fracture | ✓ | ~ | ✓ | ✓ | ✓ | ✓ | ✓ | ✓ |
| Veneer fracture/chipping | ✓ | ✓ | ✓ | ✓ | ✓ | ✓ | ✓ | ✓ |
| Screw loosening | — | ✓ | ✓ | ✓ | ✓ | ✓ | ✓ | — |
| Screw fracture | — | — | ✓ | ~ | — | — | ✓ | — |
| Delamination | — | — | ✓ | — | — | — | — | — |
| Prosthesis remake | ✓ | ✓ | ✓ | ✓ | ✓ | ✓ | ✓ | ✓ |
| Biological complications | — | ✓ | ✓ | ~ | ~ | — | ~ | ~ |
| Peri-implantitis | — | ✓ | ✓ | ~ | ✓ | — | — | — |
| Marginal bone loss | — | ✓ | ✓ | ✓ | — | — | ~ | ~ |
| *SECONDARY OUTCOMES* |  |  |  |  |  |  |  |  |
| Implant survival<br>rate | ✓ | ✓ | ✓ | ✓ | ✓ | ✓ | ✓ | ✓ |
| Plaque index | — | ✓ | — | ✓ | — | — | — | ✓ |
| Bleeding on probing | — | ✓ | — | ✓ | — | — | — | ✓ |
| Probing depth | — | ✓ | ~ | ✓ | — | — | — | ✓ |
| Patient satisfaction | — | — | ✓ | — | — | — | ✓ | — |
| OHIP / OHRQoL | — | — | ✓ | — | — | — | ✓ | — |
| Aesthetic<br>satisfaction | — | — | ✓ | — | — | — | ✓ | — |
| *COST DATA* |  |  |  |  |  |  |  |  |
| Initial fabrication<br>cost | — | — | — | — | ✓ | — | — | — |
| Maintenance/repair<br>cost | — | — | — | — | ✓ | — | — | — |
| Total cost over<br>follow-up | — | — | — | — | ✓ | — | — | — |

✓ = reported; — = not reported; ~ = partially reported.

### 3.3 Material tier classification

All 8 studies were classified as Tier 1. Only 2 studies did not report the specific metal alloy of the prosthetic framework (Barootchi, 2020 and Box, 2018) [16, 19]. Neither of these studies did not specify the veneering material used, but both featured monolithic zirconia prostheses with both arms using CAD/CAM, therefore it is possible that both could be assigned to Tier 3 given perfect information. Box, 2018 replied to confirm titanium was the alloy used. Only one study (Tang, 2023) specified titanium as the framework material in the manuscript [43]. Three studies (Barootchi, 2020; Box, 2018; Capparé, 2021) did not specify the type of acrylic used [16, 17, 19]. Four studies explicitly used monolithic zirconia (Sabău, 2025; Durrani, 2025; Ayna, 2024; Box, 2018), with two using zirconia prosthetics on a titanium bar (Sabău, 2025; Durrani, 2025) [19, 40, 41, 42]. These results prevented the pre-specified Tier 2 and Tier 3 sensitivity analysis, and confirmed that single-arm studies will be used for building the CEA model.

### 3.4 Risk of bias

The lowest score given was 4/9, (Box, 2018) [19, 42]. It characterised by a small sample size, thorough exclusion criteria (excluded smokers, bruxers, diabetics) and unadjusted analysis. Durrani, 2025 had the smallest sample size (*n*_patients_ = 10) and scored 5/9, making it highly susceptible to bias and particularly imprecision [41]. Non-blinded outcome assessment was found across the studies but is expected with retrospective cohort studies. Tang, 2023 scored the highest (8/9), failing to report the follow-up percentage [43]. The majority of studies scored at the higher end of moderate quality. Adequacy of follow-up was the weakest category due to the nature of the study design, with many not reporting the specific proportion lost to follow-up, which combined with small sample sizes and single clinic studies results in high risk of attrition bias. Full scores are in Table 2.

### 3.5 Primary outcomes

#### 3.5.1 Prosthesis survival

Across both zirconia and metal-acrylic arms prosthesis survival was very high (Table 5). Four studies reported 100% survival for both arms [17, 40, 41, 42]. Our point estimates suggest the zirconia arms were weakly more favourable, however, all CIs for the RRs cross unity and therefore there is no statistically significant difference between the arms. The lowest metal framework prostheses survival rate was 83% (*±*11.1) at five-years which was from an unspecified alloy and acrylic combination [16]. Tartaglia, 2016 recorded the lowest zirconia prostheses survival rate (88.9%) which did also feature the largest prostheses sample size [20]. Figure 2 summarises the primary outcomes according to risk of bias scores and statistical significance.

**Table 5:** Summary of findings by outcome domain (Tier 1 analysis)

| Outcome | N studies,<br>participants,<br>prostheses | Effect estimate (RR ZrO<sub>2</sub> vs MF) | Direction | GRADE<br>certainty |
| --- | --- | --- | --- | --- |
| *PRIMARY OUTCOMES* |  |  |  |  |
| Prosthesis<br>survival | 5 studies | Tang, 2023: RR 0.72 (0.07–7.38) | Weakly favours | ⊕ ○ ○ ○ |
|  | (*n* = 266 | Barootchi, 2020: RR 0.40 (0.09–1.78) | ZrO<sub>2</sub> (NS) | Very low |
|  | participants; 396<br>prostheses) | Tartaglia, 2015: RR 0.69 (0.28–1.71) |  |  |
| Technical<br>complications<br>(overall) | 6 studies | Heterogeneous definitions | No consistent | ⊕ ○ ○ ○ |
|  | (*n* = 273; 427<br>prostheses) | Tang, 2023: reported OR 1.07 (0.32–3.59)<br>for veneer chipping (*P* = 0.911) | difference | Very low |

Table 5 continued from previous page
| Outcome | N studies,<br>participants,<br>prostheses | Effect estimate (RR ZrO<sub>2</sub> vs MF) | Direction | GRADE<br>certainty |
| --- | --- | --- | --- | --- |
| Framework<br>fracture | 5 studies<br>(*n* = 216; 318<br>prostheses) | Tang, 2023: RR 1.44 (0.10–21.62)<br>Barootchi, 2020: RR 0.83 (0.21–3.23)<br>Ayna, 2024; Durrani, 2025; Capparé, 2021: 0<br>events both arms | No difference | ⊕ ○ ○ ○<br>Very low |
| Veneer<br>fracture/chipping | 5 studies<br>(*n* = 199; 281<br>prostheses) | Tang, 2023: RR 0.96 (0.50–1.87)<br>Barootchi, 2020: RR 0.46 (0.30–0.71)<br>Ayna, 2024: RR 1.00 (0.47–2.15)<br>Durrani, 2025: RR 0.14 (0.01–2.49) | Weakly favours<br>ZrO<sub>2</sub> (NS) | ⊕ ○ ○ ○<br>Very low |

Table 5 continued from previous page
| Outcome | N studies,<br>participants,<br>prostheses | Effect estimate (RR ZrO<sub>2</sub> vs MF) | Direction | GRADE<br>certainty |
| --- | --- | --- | --- | --- |
| Screw loosening | 4 studies<br>(*n* = 126; 178<br>prostheses) | Ayna, 2024: RR 0.20 (0.01–3.84)<br>Barootchi, 2020: RR 1.11 (0.32–3.80)<br>Durrani, 2025: RR 0.23 (0.01–4.93)<br>Tang, 2023: 0 events both arms | No difference<br>(NS) | ⊕ ○ ○ ○<br>Very low |
| Biological<br>complications<br>(overall) | 4 studies<br>(*n* = 126; 188<br>prostheses) | Peri-implantitis (implant level):<br>Tang, 2023: RR 0.49 (0.10–2.36)<br>Barootchi, 2020: RR 1.30 (0.91–1.86) | No consistent<br>difference | ⊕ ○ ○ ○<br>Very low |
| ***SECONDARY OUTCOMES*** |  |  |  |  |

Table 5 continued from previous page
| Outcome | N studies,<br>participants,<br>prostheses | Effect estimate (RR ZrO<sub>2</sub> vs MF) | Direction | GRADE<br>certainty |
| --- | --- | --- | --- | --- |
| Implant survival | 6 studies<br>(*n* = 259; ~826<br>implants) | Tang, 2023: RR 7.28 (0.35–150.23) [2 events<br>vs 0 events]<br>Barootchi, 2020: RR 0.76 (0.44–1.30) (est.)<br>Ayna, 2024; Durrani, 2025; Capparé, 2021:<br>100% both arms<br>Tartaglia, 2015: 94% overall (not separated) | No difference | ⊕ ○ ○ ○<br>Very low |

Table 5 continued from previous page
| Outcome | N studies,<br>participants,<br>prostheses | Effect estimate (RR ZrO<sub>2</sub> vs MF) | Direction | GRADE<br>certainty |
| --- | --- | --- | --- | --- |
| Marginal bone<br>loss (mm) | 3 studies ($n = 70$;<br>94 prostheses) | MD (ZrO<sub>2</sub> minus MF):<br>Ayna, 2024 (straight): MD $-0.93$ ($-1.01$ to<br>$-0.85$)<br>Ayna, 2024 (tilted): MD $-1.10$ ($-1.20$ to<br>$-1.00$)<br>Tang, 2023: MD $-0.17$ ($-0.41$ to $0.07$)<br>Durrani, 2025: MD $-1.10$ ($-1.32$ to $-0.88$) | Favours ZrO<sub>2</sub><br>(consistent) | $\oplus \circ \circ \circ$<br>Very low |

Table 5 continued from previous page
| Outcome | N studies,<br>participants,<br>prostheses | Effect estimate (RR ZrO<sub>2</sub> vs MF) | Direction | GRADE<br>certainty |
| --- | --- | --- | --- | --- |
| Plaque index | 3 studies ($n = 90$;<br>100 prostheses) | Capparé, 2021 (FMPS, site-level): RR 0.70<br>(0.61–0.81)<br>Ayna, 2024: significant ($P < 0.001$;<br>quantitative data NR)<br>Durrani, 2025: qualitatively lower | Favours ZrO<sub>2</sub><br>(consistent) | $\oplus \circ \circ \circ$<br>Very low |
| Bleeding on<br>probing | 3 studies ($n = 90$;<br>100 prostheses) | Capparé, 2021 (FMPS, site-level): RR 0.39<br>(0.27–0.58)<br>Ayna, 2024: significantly lower ($P < 0.001$;<br>quantitative data NR)<br>Durrani, 2025: qualitatively lower | Favours ZrO<sub>2</sub><br>(consistent) | $\oplus \circ \circ \circ$<br>Very low |

Table 5 continued from previous page
| Outcome | N studies,<br>participants,<br>prostheses | Effect estimate (RR ZrO<sub>2</sub> vs MF) | Direction | GRADE<br>certainty |
| --- | --- | --- | --- | --- |
| Probing depth | 2 studies ($n = 60$;<br>50 prostheses) | Capparé, 2021: MD $-0.22$ ($-0.65$ to $0.21$)<br>Durrani, 2025: ZrO<sub>2</sub> 3 mm vs MF 3–4 mm<br>(SD NR; CI not estimable) | Weakly favours<br>ZrO<sub>2</sub> (NS) | $\oplus \circ \circ \circ$<br>Very low |
| Patient<br>satisfaction /<br>OHRQoL | 2 studies ($n = 67$;<br>93 prostheses) | Box, 2018: OHIP-49 7–29 across all groups;<br>$P = 0.16$ (no between-group difference)<br>Tang, 2023: >80% very satisfied (combined<br>arms; no between-group test reported) | No difference | $\oplus \circ \circ \circ$<br>Very low |

Table 5 continued from previous page
| Outcome | N studies,<br>participants,<br>prostheses | Effect estimate (RR ZrO<sub>2</sub> vs MF) | Direction | GRADE<br>certainty |
| --- | --- | --- | --- | --- |
| Cost data | 1 study ($n = 56$;<br>74 prostheses) | Barootchi, 2020:<br>Initial cost: ZrO<sub>2</sub> \$20,518 vs MF \$12,024;<br>MD +\$8,494 (CI not estimable; aggregate<br>data only)<br>Maintenance: ZrO<sub>2</sub> \$2,185 vs MF \$2,041;<br>MD +\$144 | Higher costs for<br>ZrO<sub>2</sub> | $\oplus \circ \circ \circ$<br>Very low |

Abbreviations: RR, risk ratio; MD, mean difference; CI, confidence interval; K–M, Kaplan–Meier; NS, not statistically significant; NR, not reported; FMPS, full-mouth plaque score; FMBS, full-mouth bleeding score; OHRQoL, oral health-related quality of life; est., estimated from aggregate data.

**Table 6:** GRADE evidence profile for included outcomes.

| Outcome | N | Risk of bias | Inconsistency | Indirectness | Imprecision | Pub. bias | Large effect? | Dose-resp.? | Residual conf.? | GRADE |
| --- | --- | --- | --- | --- | --- | --- | --- | --- | --- | --- |
| Prosthesis survival | 5 | S | NS | NS | S | U | No | No | No | ⊕ ○ ○ ○ |
| Tech. comp. (overall) | 6 | S | S | S | S | U | No | No | No | ⊕ ○ ○ ○ |
| Framework fracture | 5 | S | NS | S | VS | U | No | No | No | ⊕ ○ ○ ○ |
| Veneer fract./chipping | 6 | S | S | S | S | U | No | No | No | ⊕ ○ ○ ○ |
| Biological comp. | 4 | S | S | S | S | U | No | No | No | ⊕ ○ ○ ○ |
| Implant survival | 6 | S | NS | NS | S | U | No | No | No | ⊕ ○ ○ ○ |
| Plaque index | 4 | S | S | S | S | U | No | No | No | ⊕ ○ ○ ○ |
| Bleeding on probing | 4 | S | S | S | S | U | No | No | No | ⊕ ○ ○ ○ |
| Probing depth | 4 | S | S | S | S | U | No | No | No | ⊕ ○ ○ ○ |
| Patient sat. / OHRQoL | 2 | S | S | S | S | U | No | No | No | ⊕ ○ ○ ○ |

*S = Serious; NS = Not serious; VS = Very serious; U = Undetected. ⊕ ○ ○ ○ = Very low.*
*Non-randomised evidence starts at low certainty.*

**Table 7:** Model input parameters for cost-effectiveness analysis.

| Parameter | Base case | Range<br>(low–high) | Distribution Source |
| --- | --- | --- | --- |
| *Cost Inputs* |  |  |  |
| Placement Surgery | Ti-PMMA = £17,838;<br>Zirconia = £24,635 | £16,000 to<br>£35,000 | Gamma |
| Annual maintenance | £336 | £150 to £400 | Gamma |
| Minor repair (per event) | Ti-PMMA = £436;<br>Zirconia = £586 | £150 to £800 | Gamma |
| Major remake | Ti-PMMA = £2,923;<br>Zirconia = £25,423 | £5,000 to £18,000 | Gamma |
| Implant placement | £2,896 | £1,500 to £4,000 | Gamma |
| Prosthesis failure ongoing<br>annual cost | £566 | £300 to £800 | Gamma |
| *Utility/Disutility Parameters* |  |  |  |
| Disutility: minor<br>complication (S2) | −0.050 | −0.01 to −0.100 | Beta |
| Duration: minor<br>complication (frac. year) | 0.038 | 0.019 to 0.077 | Fixed |
| Disutility: major<br>complication (S3) | −0.150 | −0.100 to −0.250 | Beta |
| Duration: major<br>complications (frac. year) | 0.130 | 0.077 to 0.230 | Fixed |
| Disutility: implant loss (S4) | −0.200 | −0.100 to −0.300 | Beta |
| Duration: implant loss (frac.<br>year) | 0.250 | 0.125 to 0.500 | Fixed |
| Prosthesis failure multiplier<br>(S5) | 0.100 | 0.050 to 0.150 | Beta |

*All costs in 2026 UK pounds sterling and from the patient's perspective.*

#### 3.5.2 Technical complications

Technical complications were common across both arms but due to varying definitions we were unable to determine single-pooled estimates. Three studies reported zero framework fractures for both arms [17, 41, 42]. Although, framework fractures were generally uncommon across both arms. The majority of reported technical complications came from veneer fracture and/or chipping. Of the studies that reported veneer fracture and/or chipping by prostheses sub-type, there was only one instance where zero was reported (Durrani, 2025; monolithic zirconia framework layered with lithium disilicate on titanium bar) [41]. In the study with the longest follow-up Barootchi, 2020 reported that the zirconia arm showed a statistically significant reduction in veneer fracture/chipping, although all other studies reporting showed no difference [16]. Our analysis and conclusions here are limited by the mixture of ceramic and acrylic used in the comparator arm. Screw loosening was either not reported or very low across both arms in all studies. Multiple studies report that the minor complications in acrylic veneering were fixed chair-side and did not require complex remaking [40, 42].

#### 3.5.3 Biological complications

Biological complications results were conflicting, with higher mucositis and peri-implantitis prevalence reported in the zirconia arm under Barootchi, 2020, but the opposite was reported by Tang, 2023 [16, 43]. Soft tissue complications were fairly common for both arms in studies that reported them, although reported narratively. No meaningful or reliable difference between the groups could be established. Only one study reported the effect of the prostheses on the opposing arch (Box, 2018) [19]. Both the monolithic zirconia and porcelain-veneered zirconia had a higher average number of complications to opposing arch per prosthesis, however, we mark this as an evidence gap due to it being unreported by other studies.

### 3.6 Secondary outcomes

#### 3.6.1 Implant survival

Implant survival rates across both arms were near identical. Four studies (Sabău, 2025; Durrani, 2025; Ayna, 2024; Capparé, 2021) reported 100% for both arms, with Tang, 2023 reporting 100% for titanium-ceramic and 98.2% for zirconia-ceramic [17, 40, 41, 42, 43]. Only Box, 2018 did not report implant survival, however, the focus of that paper was more on patient-reported and final prosthetic outcomes [19]. The lowest implant survival rate was reported by Barootchi, 2020 in their metal-acrylic arm (87.3%; zirconia = 90.5%). The study was on both immediate and delayed loaded prosthetics, but final results were not aggregated by this distinction and the results not statistical significant [16]. Implant survival was also the only secondary outcome that was reported in this study.

#### 3.6.2 Peri-implant clinical parameters

Across all recorded parameters (plaque index, bleeding on probing and probing depth), the zirconia arms outperformed the metal framework arms. Only Capparé, 2021 and Ayna, 2024 used statistical significant testing to determine a meaningful statistical difference between the two arms [17, 42]. When reported qualitatively, Durrani, 2025 reported plaque accumulation and bleeding on probing to be lower in the zirconia arm, although this prosthetic did use a titanium bar and lithium disilicate layering and featured a very small sample size across both arms (*n*_prostheses_ = 20) [41].

#### 3.6.3 Patient-reported outcome measures

PROMs were sparsely reported, with only two studies directly asking patients for their satisfaction level (Tang, 2023; Box, 2018), and only one used the OHIP-49 questionnaire (Box, 2018) [19, 43]. Difference between the groups were not statistically significant using OHQoL scores, but both arms scored excellent (*P* = .16). Monolithic zirconia scored the lowest OHIP-49 score, but veneered zirconia the highest. They also found that number of complications was not a predictor of OHQoL. We highlight the lack of reported PROMs as an evidence gap.

### 3.7 Subgroup and sensitivity analyses

We were unable to conduct any pre-specified analyses due to the variability in evidence and heterogeneity in definitions used.

### 3.8 Certainty of evidence

Table 6 provides the GRADE evidence profile. All studies were non-randomised comparative cohort studies which means all evidence started at a low certainty level. Due to the retrospective study design, potential confounding individual characteristics and variability in methodological quality the certainty for several outcomes was downgraded due to the risk of bias. We saw heterogeneity in outcome definitions, such as peri-implant soft tissue parameters and veneer fracture/chipping, across studies which led to downgrades for inconsistency and indirectness. Small sample sizes and few outcomes reported resulted in a downgrade for imprecision. Overall, the overall certainty of evidence was decided to be very low. Our finding is consistent with oral health systematic reviews [44]. Full explanation for why certain studies were excluded from GRADE evidence outcomes can be found in the Supplementary Material (Table S8).

**Part 2: Cost-Effectiveness Analysis**

### 3.9 Model parameters

The direct comparative literature was sparse for the two dominant prosthetic approaches, therefore, we have used the most suitable single-arm data in the broader literature to model this comparison that clinical studies are yet to address. This absence of evidence supports the rationale for modelling. The search strings used for the parameter search are found in Supplementary Material Table S8, while literature previously found in the systematic review section was also hand searched for relevant utility values, transition probabilities and cost data. Table 7 details our final model parameters used. We utilised single-arm studies with significant follow-up and prostheses sample size to determine transition probabilities (full justification in Supplementary Material Table S7) [16, 45, 46, 47, 48, 49]. Our State 1 remain functioning probability for Ti-PMMA was calculated as 83.3% and 96.0% for zirconia. Given the available evidence, we deemed it necessary to make the probability of functioning to implant loss, implant loss to functioning and implant loss to failure the same for both arms, as evidence suggests this does not differ by prosthesis material and that failure is driven by non-material factors [16, 50]. Transition probabilities are treated as per-patient probabilities.

### 3.10 Base-case results

Total costs for both arms were dominated by the initial surgical placement cost, which was expected given the high prostheses survival rate and probability of functioning. Total discounted costs for the Ti-PMMA arm were £23,295,011 and £29,169,691 for the zirconia arm. Total QALYs were 10,259.95 for Ti-PMMA, and 10,278.58 for zirconia. This was expected given the same utility and disutility values were used for both. This resulted in an ICER of £315,417, which means that zirconia is not cost-effective at the NHS willingness-to-pay thresholds, which was expected and not entirely important given the patient-funded nature of dental care. Zirconia prostheses were both more costly, with a per-patient net monetary benefit of *−*£5,502.18 at the £20,000 threshold and *−*£5,315.93 at the £30,000 threshold. Total discounted per-patient costs over a 20-year horizon for full-jaw Ti-PMMA prosthesis were calculated to be £23,295.01, and £29,169.69 for zirconia.

### 3.11 Sensitivity analyses

#### 3.11.1 Deterministic sensitivity analysis

Varying the State 1 to State 3 transition probability, functioning to major complication, for both arms caused the greatest difference in the final ICER, followed by changing the initial placement cost for both Ti-PMMA and Zirconia (Figure 4a). The majority of parameters had no significant effect on the final ICER, including the minor and major remake cost and disutility parameters. No parameter variation resulted in zirconia becoming cost effective at either the £20,000 or £30,000 threshold.

**Figure 4:**
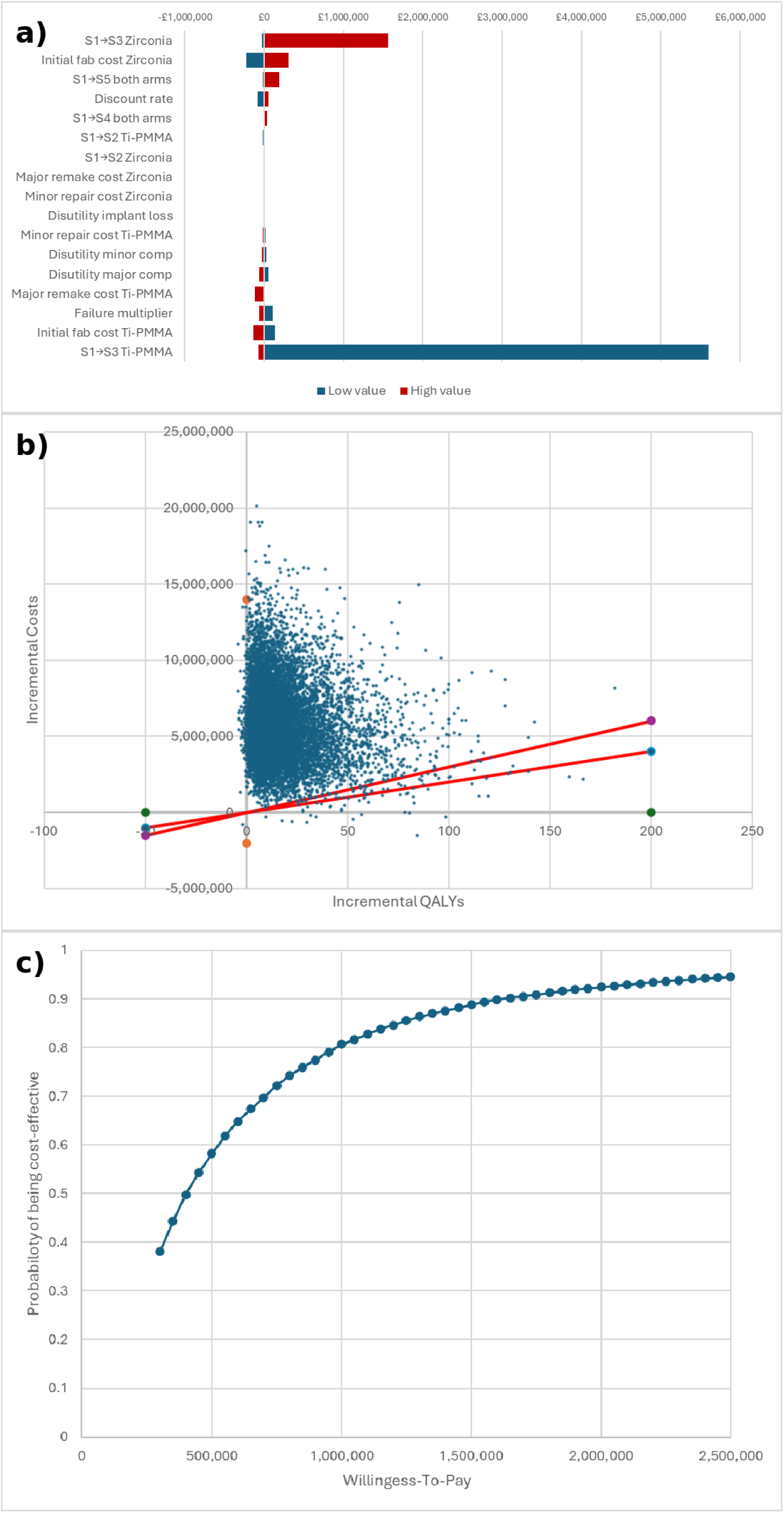
*(a) Tornado diagram, (b) Cost-effectiveness plane, (c) Cost-effectiveness acceptability curve.* Cost-effectiveness analysis results. (a) Tornado diagram showing one-way deterministic sensitivity analysis; parameters ranked by influence on ICER. (b) Cost-effectiveness plane from 10,000 Monte Carlo simulations; dashed line represents £20,000/QALY threshold. (c) Cost-effectiveness acceptability curve across willingness-to-pay thresholds.

#### 3.11.2 Probabilistic sensitivity analysis

Our PSA results reveal that at a threshold of £20,000, zirconia was cost-effective in 0.50% of the iterations, whereas Ti-PMMA was cost-effective in 99.50% of iterations (Figure 4b). For the majority of PSA iterations, zirconia prostheses was both more effective and more costly than Ti-PMMA. The probabilistic ICER (ratio of the mean incremental costs to mean incremental QALYs) of £320,954.22 was in line with our base case. Average per-patient costs for zirconia (£29,159.2, CI = £22,625.08 to £36,966.86) and Ti-PMMA (£23,281.45, CI = £18,368.26 to £28,940.99) were consistent with the base run.

#### Scenario analyses

Reducing the discount rate to 1% resulted in a 26% reduction in the ICER, to £232,707.27, while increasing it to 5% increased the ICER by 18% to £372,014.14. This pattern was expected given the majority of upfront costs and how the prostheses benefit’s accrue over the long term. We tested a copayment system at 25%, 50% and 75% to simulate a situation where patient costs are partially covered by either insurance or the NHS. A 25% copayment means the patient only pays 75% of the total direct cost. The copayment was applied to both prostheses simultaneously. This applied only to the direct healthcare costs of initial placement surgery, and did not affect travel or productivity costs. At 25%; the ICER dropped to £224,192 (per-patient costs: Ti-PMMA = £18,920.84; Zirconia = £23,096.44), 50%; ICER at £132,966 (per-patient costs: Ti-PMMA = £14,546.68; Zirconia = £17,023.19); and 75%; ICER at £41,741 (per-patient costs: Ti-PMMA = £10,172.51; Zirconia = £10,949.94).

## 4. DISCUSSION

### 4.1 Summary of main findings

Across all arms, prosthetic- and patient-level outcomes were overwhelming positive regardless of the framework or veneering material used. However, this systematic review highlights the low certainty of the evidence available. All studies found high prostheses and implant survival across both material approaches but with no statistically significant difference for any of the primary outcomes measured. All evidence was graded at low uncertainty, primarily due to small sample size, no RCTs and inconsistent material reporting which resulted in imprecision and risk of bias (Table 6). Zirconia emerged as favourable for peri-implant parameters, although no studies measured the effect of cleansing pockets and how their effect on plaque accumulation varied with the material used, which reflects clinical reality as patients returning for check-ups will often receive cleaning. Technical complications, such as minor and major chipping, resulted in more complex to interpret results. Acrylic veneering was often found to suffer from small chipping and fractures, however, these were routinely repaired chairside without costly lab intervention. Zirconia prostheses suffered from less frequent minor complications, but they tended to require a greater level of intervention. Minor chipping and fractures were reported per event, without any details of the severity or size of each. We highlight the high success rate and patient satisfaction with ISFCDPs, regardless of the material used. Regardless, the lack of consistent PROMs reporting needs to be highlighted as an evidence gap in this literature, with factors such as patient speech, pain in the mouth and satisfaction with the prosthetic remaining largely unreported. Our integrated CEA found that there is approximately a £6,000 per-patient premium attached to monolithic zirconia for only a marginal gain in QALYs. Our very high ICER (£315,417) is almost purely the result of the cost differential. We provide a novel open-source CEA model for use that can be modified as new evidence and estimates emerge. We have used monolithic zirconia and Ti-PMMA as our comparators of choice, however, the flexibility of the model allows values of other zirconia and metal-acrylic combinations to be easily integrated in assuming the data is available.

### 4.2 Comparison with existing literature

Our tiered approach was designed to provide a methodological contribution for the area, testing whether conclusions changed with increasing metal alloy specificity. However, the vast heterogeneity in how prostheses are reported and the materials used limited its application. All eight studies were categorised as Tier 1, and only two had confirmed the alloy as titanium, and multiple not specifying the type of acrylic.

The limitations we find in the reported studies are very similar to previous reviews. Short follow-up times and a low number of treated patients has been previously cited as a characteristic of studies evaluating zirconia ISFCDPs, partially due to their relatively recent emergence as a treatment option [7]. This was highlighted in a review of single-arm studies, and we found the same when evaluating two-armed studies. Delucchi et al. grouped all metal alloys together without addressing the veneering material, which makes comparisons hard [8]. The very high prostheses and implant survival rate we found are established in the single-arm reviews, although our within-study comparisons found that the survival advantage for zirconia diminishes when confidence intervals are calculated. We hoped to test the biocompatibility and corrosion differences between titanium and cobalt-chromium frameworks through our tiered approach, however the available clinical literature did not allow this [8]. Both Cinquini et al., Delucchi et al. and Al-Tarawneh et al. noted material heterogeneity as an issue, but neither attempted to classify it [11]. Only Barootchi et al. included cost data but their analysis was only descriptive and without an attempt to model on a real-world cohort. Their US-based data demonstrated a similar pattern to our UK-survey approach. They concluded that there was higher initial and maintenance costs for zirconia, yet no significant increase in benefits [16]. Zhurakivska et al. has previously modelled the cost-effectiveness for mandibular edentulism rehabilitation comparing variations of traditional dentures with fixed-implant dentures, not comparing by material [51]. To our knowledge, this is the first published CEA to compare two different material approaches for ISFCDPs. We have designed this model to be updated based on the materials that want to be compared.

### 4.3 Clinical implications

Any direct clinical limitations need to be treated with caution due to the lack of evidence certainty, and absence of RCTs directly testing zirconia and metal framework-acrylic prostheses. Both prosthetic designs result in high survival success rates, with no statistically significant difference detected for any of the primary outcomes. The primary clinical result is that zirconia prostheses showed slightly more favourable peri-implant outcomes, but acrylic veneering chipping was easier and quicker to fix than zirconia complications. The presence of strict exclusion criteria in some studies also limits clinical implications. Three studies featured exclusion criteria that prevented analysis on prostheses in individuals with characteristics typical of those who seek full-jaw rehabilitation [17, 20, 42]. This included having bruxismus, poor motivation to accept an oral hygiene maintenance program, uncontrolled diabetes and being a smoker. Furthermore, only two studies reported PROMs, which although were overwhelming positive, still does not provide enough evidence to determine overall patient satisfaction and these results need to be treated with caution [19, 43].

### 4.4 Economic implications

Our results determine that the additional cost of zirconia does not justify the additional patient benefits as they are only marginal when compared to Ti-PMMA. A large majority of costs, across both arms, related to full-jaw prosthetics was estimated to come from the initial placement surgery. This is owed to the high success rate which overall minimises repair and remake costs. Our estimated ICER of £315,417 is far more than the NICE suggested willingness-to-pay thresholds. Given the out-of-pocket nature of dental care and its position in the private UK market, we determined to treat comparisons to this threshold cautiously. The £20,000 to £30,000 per QALY threshold was designed to be used from a public healthcare payers perspective under tight resource allocation. While our conclusions related to the threshold are true and important, we believe that per-patient cost analysis is more decision-relevant. Over the 20-year horizon the total cost difference between the Ti-PMMA and monolithic zirconia approach was estimated to be approximately £5,875. This purchased a very small QALY gain of 0.019. We took the patient perspective due to its relevance in the UK setting. Therefore, the individual patient decision should be determined on whether that additional cost justifies the slightly smaller complication profile and the peri-implant health advantages found in our systematic review. However, even these advantages were deemed not statistically significant with RRs and CIs crossing unity. Our PSA confirmed this conclusion, with net monetary benefit consistently negative for zirconia across both NICE thresholds (Supplementary Material Table S9). The copayment scenario was exploratory analysis of a hypothetical partial subsidy for advanced dental care. Even when a 75% subsidy was provided to the patient the ICER remained above the thresholds at £41,741. This did substantially reduce the per-patient costs for both arms (Ti-PMMA = £10,172.51; Zirconia = £10,949.94), but further willingness-to-pay research needs to determine what effect this would have on the demand and financial accessibility of full-jaw rehabilitation. Our one-way sensitivity analysis (Figure 4a) determined that the functioning-to-major complication transition probability was the most influential parameter for both arms. This means that if studies with a greater number of patients and longer follow-ups showed a larger statistically significant difference in major complications rates between materials, the ICER could shift significantly.

### 4.5 Strengths and limitations

The main limitations of our analysis are due to the type of studies that were available for review. This resulted in a small and heterogeneous evidence base with most outcome differences not being statistically significant. The follow-up times of studies included was markedly small compared to the expected prosthesis lifespan, and the lack of prospective controlled studies meant confounding variables were often not considered. As a result, all evidence was given a GRADE certainty of very low. Our cost assumptions do not include loan and financed options which will increase the total cost for a patient. There will be heterogeneity in the fine details of the prosthetics used, including the number of teeth and depth of smile, which will affect patient satisfaction and the quality of the prosthetic. Some studies reported that bone grafting was used for some of the implants, whereas others made no mention. This smaller details are hard to account for but may still influence outcomes in the CEA. The main strength of our study is that, to our knowledge, this is the first head-to-head comparison of different materials used in ISFCDPs with integrated CEA. We have used transparent methods compliant to PRISMA and SWiM methodologies.

### 4.6 Implications for future research

Prospective studies and adequately powered RCTs with transparent alloy specification are needed to assess and compare the two dominant full-jaw treatment options. Follow-up periods need to be a minimum of 5 years and there needs to be clarity in pre-specifying the definitions of minor and major complications. Multi-centre designs need to be considered, as all eight of the included studies were from single-centres. Most importantly, the reporting of framework and veneering materials needs to be mandatory. We believe the largest evidence gap our systematic review has located is the lack of PROMs in the evaluation of full-jaw rehabilitation. Given its convenience to both researchers and patients, we believe standardised measures such as OHIP-49 need to be used more actively. This would help to inform more accurate CEA by allowing models to step away from more generic utility value sets and more condition-specific utility values that better capture the effects of ISFCDPs.

## 5. CONCLUSIONS

*Both zirconia and metal-framework veneered ISFCDPs resulted in high prosthesis and implant survival, but we determined no statistically significant difference in recorded primary clinical outcomes. The evidence available is overall very weak, given the small sample sizes, short follow-up and heterogenous outcome definitions. The approximately £5,875 per-patient cost premium for monolithic zirconia produced very small per-patient QALY gains (0.019). We summarise the patient choice as a trade-off between fewer but more consequential zirconia complications and more frequent but chair side repairable acrylic complications*.

## DECLARATIONS

### Author contributions

EC: Methodology, Formal analysis, Investigation, Data curation, Writing and Visualisation. NP: Reviewing selected literature, Validation, Investigation, reviewing and editing final manuscript. RV: Study conception and design, Review and edits of final manuscript.

### Data availability

All data extracted from included studies are presented in the manuscript and supplementary materials. The complete search strategy, extraction forms, and Python code for meta-analysis and Excel used for economic modelling are available on OSF [10.17605/OS-FJO/YHM46] and GitHub.

## Data Availability

All data extracted from included studies are presented in the manuscript and supplementary materials.

## Acknowledgements

N/a

## Ethical approval

Not required. This study is a systematic review of published literature and does not involve primary data collection from human participants.

