## Supplementary File for "Zirconia-based versus metal-framework veneered complete-arch implant-supported fixed dental prostheses: a systematic review of comparative clinical studies with integrated cost-effectiveness analysis"

**Supplementary Material**

Supplementary Table S1 – PRISMA checklist

| **Section and Topic** | **Item #** | **Checklist item** | **Location where item is reported** |
| --- | --- | --- | --- |
| **TITLE** | | |  |
| Title | 1 | Identify the report as a systematic review. | Title page |
| **ABSTRACT** | | |  |
| Abstract | 2 | See the PRISMA 2020 for Abstracts checklist. | Page 1 |
| **INTRODUCTION** | | |  |
| Rationale | 3 | Describe the rationale for the review in the context of existing knowledge. | Page 3 |
| Objectives | 4 | Provide an explicit statement of the objective(s) or question(s) the review addresses. | Page 4 |
| **METHODS** | | |  |
| Eligibility criteria | 5 | Specify the inclusion and exclusion criteria for the review and how studies were grouped for the syntheses. | Page 5 |
| Information sources | 6 | Specify all databases, registers, websites, organisations, reference lists and other sources searched or consulted to identify studies. Specify the date when each source was last searched or consulted. | Page 6 |
| Search strategy | 7 | Present the full search strategies for all databases, registers and websites, including any filters and limits used. | Abstract |
| Selection process | 8 | Specify the methods used to decide whether a study met the inclusion criteria of the review, including how many reviewers screened each record and each report retrieved, whether they worked independently, and if applicable, details of automation tools used in the process. | Page 7 |
| Data collection process | 9 | Specify the methods used to collect data from reports, including how many reviewers collected data from each report, whether they worked independently, any processes for obtaining or confirming data from study investigators, and if applicable, details of automation tools used in the process. | Page 7 |
| Data items | 10a | List and define all outcomes for which data were sought. Specify whether all results that were compatible with each outcome domain in each study were sought (e.g. for all measures, time points, analyses), and if not, the methods used to decide which results to collect. | Page 7-8 |
|  | 10b | List and define all other variables for which data were sought (e.g. participant and intervention characteristics, funding sources). Describe any assumptions made about any missing or unclear information. | N/a |
| Study risk of bias assessment | 11 | Specify the methods used to assess risk of bias in the included studies, including details of the tool(s) used, how many reviewers assessed each study and whether they worked independently, and if applicable, details of automation tools used in the process. | Page 9 |
| Effect measures | 12 | Specify for each outcome the effect measure(s) (e.g. risk ratio, mean difference) used in the synthesis or presentation of results. | Page 9 |
| Synthesis methods | 13a | Describe the processes used to decide which studies were eligible for each synthesis (e.g. tabulating the study intervention characteristics and comparing against the planned groups for each synthesis (item #5)). | Page 6-7 |
|  | 13b | Describe any methods required to prepare the data for presentation or synthesis, such as handling of missing summary statistics, or data conversions. | Page7 |
|  | 13c | Describe any methods used to tabulate or visually display results of individual studies and syntheses. | Page 8-10 |
|  | 13d | Describe any methods used to synthesize results and provide a rationale for the choice(s). If meta-analysis was performed, describe the model(s), method(s) to identify the presence and extent of statistical heterogeneity, and software package(s) used. | Page 9 |
|  | 13e | Describe any methods used to explore possible causes of heterogeneity among study results (e.g. subgroup analysis, meta-regression). | Page 7-8 |
|  | 13f | Describe any sensitivity analyses conducted to assess robustness of the synthesized results. | Page 7-8 |
| Reporting bias assessment | 14 | Describe any methods used to assess risk of bias due to missing results in a synthesis (arising from reporting biases). | N/a |
| Certainty assessment | 15 | Describe any methods used to assess certainty (or confidence) in the body of evidence for an outcome. | Page 10 |
| **RESULTS** | | |  |
| Study selection | 16a | Describe the results of the search and selection process, from the number of records identified in the search to the number of studies included in the review, ideally using a flow diagram. | Page 13-14 |
|  | 16b | Cite studies that might appear to meet the inclusion criteria, but which were excluded, and explain why they were excluded. | Page 13 |
| Study characteristics | 17 | Cite each included study and present its characteristics. | Page 19-20 |
| Risk of bias in studies | 18 | Present assessments of risk of bias for each included study. | Page 15-16 |
| Results of individual studies | 19 | For all outcomes, present, for each study: (a) summary statistics for each group (where appropriate) and (b) an effect estimate and its precision (e.g. confidence/credible interval), ideally using structured tables or plots. | Page 26-33 |
| Results of syntheses | 20a | For each synthesis, briefly summarise the characteristics and risk of bias among contributing studies. | Page 15-16 |
|  | 20b | Present results of all statistical syntheses conducted. If meta-analysis was done, present for each the summary estimate and its precision (e.g. confidence/credible interval) and measures of statistical heterogeneity. If comparing groups, describe the direction of the effect. | Page 26-33 |
|  | 20c | Present results of all investigations of possible causes of heterogeneity among study results. | Page 25 |
|  | 20d | Present results of all sensitivity analyses conducted to assess the robustness of the synthesized results. | Page 25 |
| Reporting biases | 21 | Present assessments of risk of bias due to missing results (arising from reporting biases) for each synthesis assessed. | N/a |
| Certainty of evidence | 22 | Present assessments of certainty (or confidence) in the body of evidence for each outcome assessed. | Page 25, Page 34 |
| **DISCUSSION** | | |  |
| Discussion | 23a | Provide a general interpretation of the results in the context of other evidence. | Page 41-42 |
|  | 23b | Discuss any limitations of the evidence included in the review. | Page 44-45 |
|  | 23c | Discuss any limitations of the review processes used. | Page 44-45 |
|  | 23d | Discuss implications of the results for practice, policy, and future research. | Page 43-45 |
| **OTHER INFORMATION** | | |  |
| Registration and protocol | 24a | Provide registration information for the review, including register name and registration number, or state that the review was not registered. | Page 2 |
|  | 24b | Indicate where the review protocol can be accessed, or state that a protocol was not prepared. | Page 2 |
|  | 24c | Describe and explain any amendments to information provided at registration or in the protocol. | N/a |
| Support | 25 | Describe sources of financial or non-financial support for the review, and the role of the funders or sponsors in the review. | Page 46 |
| Competing interests | 26 | Declare any competing interests of review authors. | Page 46 |
| Availability of data, code and other materials | 27 | Report which of the following are publicly available and where they can be found: template data collection forms; data extracted from included studies; data used for all analyses; analytic code; any other materials used in the review. | Page 47 |

Supplementary Table S2 – SWiM checklist

| **SWiM is intended to complement and be used as an extension to PRISMA** | | | |
| --- | --- | --- | --- |
| **SWiM reporting item** | **Item description** | **Page in manuscript where item is reported** | **Other*** |
| *Methods* | | | |
| **1** Grouping studies for synthesis | 1a) Provide a description of, and rationale for, the groups used in the synthesis (e.g., groupings of populations, interventions, outcomes, study design) | Section 2.8 (pp. 7–8); Section 2.10 (pp. 9); Section 3.3 (pp. 14–15) |  |
|  | 1b) Detail and provide rationale for any changes made subsequent to the protocol in the groups used in the synthesis | Section 3.3 (pp. 14–15); Section 3.7 (p. 25) |  |
| **2** Describe the standardised metric and transformation methods used | Describe the standardised metric for each outcome. Explain why the metric(s) was chosen, and describe any methods used to transform the intervention effects, as reported in the study, to the standardised metric, citing any methodological guidance consulted | Section 3.5 (pp. 16–17); Table 5 (pp. 26–33) |  |
| **3** Describe the synthesis methods | Describe and justify the methods used to synthesise the effects for each outcome when it was not possible to undertake a meta-analysis of effect estimates | Section 2.10.1 (p. 9); Section 3.5–3.6 (pp. 16–24) |  |
| **4** Criteria used to prioritise results for summary and synthesis | Where applicable, provide the criteria used, with supporting justification, to select the particular studies, or a particular study, for the main synthesis or to draw conclusions from the synthesis (e.g., based on study design, risk of bias assessments, directness in relation to the review question) | Section 2.7 (p. 7); Section 2.8 (pp. 7–8); Table 1 (p. 8) |  |
| **SWiM reporting item** | **Item description** | **Page in manuscript where item is reported** | **Other*** |
| **5** Investigation of heterogeneity in reported effects | State the method(s) used to examine heterogeneity in reported effects when it was not possible to undertake a meta-analysis of effect estimates and its extensions to investigate heterogeneity | Section 2.8 (pp. 7–8); Section 3.2 (pp. 14); Section 4.2 (pp. 42–43) |  |
| **6** Certainty of evidence | Describe the methods used to assess certainty of the synthesis findings | Section 2.11 (p. 9); Table 6 (p. 34) |  |
| **7** Data presentation methods | Describe the graphical and tabular methods used to present the effects (e.g., tables, forest plots, harvest plots).  Specify key study characteristics (e.g., study design, risk of bias) used to order the studies, in the text and any tables or graphs, clearly referencing the studies included | Table 4 (pp. 21–23); Table 5 (pp. 26–33); Figure 2 (p. 18) |  |
| *Results* | | | |
| **8** Reporting results | For each comparison and outcome, provide a description of the synthesised findings, and the certainty of the findings. Describe the result in language that is consistent with the question the synthesis addresses, and indicate which studies contribute to the synthesis | Section 3.5–3.6 (pp. 16–24); Tables 5–6 (pp. 26–34); Figure 2 (p. 18) |  |
| *Discussion* |  |  |  |
| **9** Limitations of the synthesis | Report the limitations of the synthesis methods used and/or the groupings used in the synthesis, and how these affect the conclusions that can be drawn in relation to the original review question | Section 4.5 (pp. 44–45); Section 4.1 (pp. 41–42) |  |

Supplementary Table S3 – CHEERS checklist

| **Topic** | **No.** | **Item** | **Location where item is reported** |
| --- | --- | --- | --- |
| **Title** |  |  |  |
| **Title** | 1 | Identify the study as an economic evaluation and specify the interventions being compared. | Title (p. 1): "Zirconia-based versus metal-framework veneered complete-arch implant-supported fixed dental prostheses: a systematic review of comparative clinical studies with integrated cost-effectiveness analysis" |
| **Abstract** |  |  |  |
| **Abstract** | 2 | Provide a structured summary that highlights context, key methods, results, and alternative analyses. | Abstract (pp. 1–2): Structured abstract with Objectives, Data sources, Study selection, Data extraction and synthesis, Results, and Conclusions. Reports ICER (£315,417/QALY), PSA results (0.20% cost-effective at £20,000 threshold), and per-patient cost premium (£5,875). |
| **Introduction** |  |  |  |
| **Background and objectives** | 3 | Give the context for the study, the study question, and its practical relevance for decision making in policy or practice. | Section 1 Introduction (pp. 2–4): Describes edentulism prevalence, current ISFCDP approaches, the ITI consensus statement calling for more evidence, the gap in head-to-head CEA, and states the aim to compare clinical outcomes and integrate cost-effectiveness modelling to inform patient and clinical decisions. |
| **Methods** |  |  |  |
| **Health economic analysis plan** | 4 | Indicate whether a health economic analysis plan was developed and where available. | Section 2.1 (p. 4): Protocol registered on OSF (10.17605/OSFJO/CF796). CEA methods pre-specified in protocol. CHEERS 2022 checklist followed. |
| **Study population** | 5 | Describe characteristics of the study population (such as age range, demographics, socioeconomic, or clinical characteristics). | Section 2.2.1 (p. 5): Adult patients (≥18 years) with one or both edentulous arches. Section 2.12.5 (p. 12): Model cohort starting age 60 years, mixed-sex, 1,000 individuals. Baseline utility 0.83 from Ara and Brazier regression. |
| **Setting and location** | 6 | Provide relevant contextual information that may influence findings. | Section 2.12.1–2.12.2 (pp. 10–11): UK setting, patient perspective. Costs at 2025/2026 UK pounds. NHS not covering full-jaw rehabilitation. Cost data from UK industry providers. Travel costs from HMRC mileage rates; productivity from ONS earnings data. |
| **Comparators** | 7 | Describe the interventions or strategies being compared and why chosen. | Section 2.2.2–2.2.3 (p. 5); Section 2.8 (pp. 7–8); Table 1 (p. 8): Intervention: monolithic zirconia ISFCDPs. Comparator: titanium-PMMA ISFCDPs. Tiered classification system (Tiers 1–3) developed to manage comparator heterogeneity. CEA uses Tier 1 data with single-arm literature for transition probabilities. |
| **Perspective** | 8 | State the perspective(s) adopted by the study and why chosen. | Section 2.12.2 (p. 11): Patient perspective adopted because full-jaw rehabilitation is not covered by the NHS. Includes out-of-pocket costs, travel costs, and productivity losses. |
| **Time horizon** | 9 | State the time horizon for the study and why appropriate. | Section 2.12.1 (p. 10): 20-year time horizon with 1-year cycle length, chosen to capture the expected prosthetic lifespan. |
| **Discount rate** | 10 | Report the discount rate(s) and reason chosen. | Section 2.12.2 (p. 11): 3.5% per annum for costs and outcomes, following NICE guidance. Scenario analyses at 0%, 2% (reported as 1% in results), and 5%. |
| **Selection of outcomes** | 11 | Describe what outcomes were used as the measure(s) of benefit(s) and harm(s). | Section 2.12.5 (p. 12): QALYs used as the measure of benefit. Health states include functioning prosthesis, minor and major complications, implant failure, and prosthesis failure/abandonment. |
| **Measurement of outcomes** | 12 | Describe how outcomes used to capture benefit(s) and harm(s) were measured. | Section 2.12.5 (p. 12): QALYs derived from age-sex matched UK general population norms. Baseline utility from Ara and Brazier regression equation (0.83 at age 60). Disutilities applied per health state. ONS life tables for mortality. |
| **Valuation of outcomes** | 13 | Describe the population and methods used to measure and value outcomes. | Section 2.12.5 (p. 12): No ISFCDP-specific EQ-5D values available. Baseline from Ara and Brazier UK general population norms. Disutilities: minor complication −0.05, major −0.15, implant loss −0.20, prosthesis failure multiplier 0.10. Full justification in Supplementary Table S6. |
| **Measurement and valuation of resources and costs** | 14 | Describe how costs were valued. | Section 2.12.4 (p. 11): Initial fabrication costs from real-world UK industry provider surveys (Supplementary Table S5). Maintenance and repair costs added separately. Travel costs from HMRC mileage rates. Productivity losses from ONS median hourly wage. |
| **Currency, price date, and conversion** | 15 | Report the dates of the estimated resource quantities and unit costs, plus the currency and year of conversion. | Table 7 (p. 36); Section 2.12.4 (p. 11): All costs reported in 2025/2026 UK pounds sterling (£). No currency conversion required. |
| **Rationale and description of model** | 16 | If modelling is used, describe in detail and why used. Report if the model is publicly available and where it can be accessed. | Section 2.12.1 (pp. 10): Decision tree followed by Markov cohort model with five health states plus death. 20-year horizon, 1-year cycles, 1,000 individuals. Half-cycle correction applied. Figure 3 (p. 37) shows model structure. Model publicly available on OSF (10.17605/OSFJO/YHM46) and GitHub. |
| **Analytics and assumptions** | 17 | Describe any methods for analysing or statistically transforming data, any extrapolation methods, and approaches for validating any model used. | Section 2.12.1 (p. 10); Section 2.12.3 (p. 11); Section 3.9 (pp. 34–35): Transition probabilities derived from systematic review and supplementary single-arm literature. State 1 remain-functioning probability: Ti-PMMA 83.3%, zirconia 96.0%. Implant-related transitions assumed equal across arms. Supplementary Table S7 provides full justification. |
| **Characterising heterogeneity** | 18 | Describe any methods used for estimating how the results of the study vary for subgroups. | Section 2.8 (pp. 7–8); Section 2.12.2 (p. 11): Tiered material classification tested whether conclusions changed with increasing alloy specificity. Copayment scenario analyses (25%, 50%, 75%) tested variation by payment context. No patient-level subgroup analysis due to data limitations. |
| **Characterising distributional effects** | 19 | Describe how impacts are distributed across different individuals or adjustments made to reflect priority populations. | Not directly addressed. The model assumes a homogeneous mixed-sex cohort starting at age 60. No distributional weighting or equity adjustments applied. Acknowledged as a limitation given the out-of-pocket nature of costs. |
| **Characterising uncertainty** | 20 | Describe methods to characterise any sources of uncertainty in the analysis. | Section 2.12.6 (p. 12): One-way deterministic sensitivity analysis with tornado diagram. 10,000-iteration Monte Carlo PSA. Cost-effectiveness plane and acceptability curve. Scenario analyses varying discount rate and copayment levels. All parameter ranges and distributions in Table 7 (p. 36). |
| **Approach to engagement with patients and others affected by the study** | 21 | Describe any approaches to engage patients or service recipients, the general public, communities, or stakeholders (such as clinicians or payers) in the design of the study. | Not needed. Price data was sourced from public websites. |
| **Results** |  |  |  |
| **Study parameters** | 22 | Report all analytic inputs (such as values, ranges, references) including uncertainty or distributional assumptions. | Table 7 (p. 36); Section 3.9 (pp. 34–35): All parameters reported with base case values, ranges (low–high), distributions (Gamma for costs, Beta for utilities), and sources. Supplementary Tables S5–S8 provide additional detail. |
| **Summary of main results** | 23 | Report the mean values for the main categories of costs and outcomes of interest and summarise them in the most appropriate overall measure. | Section 3.10 (pp. 37–38): Per-patient costs: Ti-PMMA £23,295, zirconia £29,170. QALYs: Ti-PMMA 10.260, zirconia 10.279 per patient. ICER £315,417/QALY. Net monetary benefit −£5,502 at £20,000 threshold, −£5,316 at £30,000 threshold. |
| **Effect of uncertainty** | 24 | Describe how uncertainty about analytic judgments, inputs, or projections affect findings. Report the effect of choice of discount rate and time horizon, if applicable. | Section 3.11 (pp. 38–39); Figure 4 (p. 40): DSA: S1→S3 transition probability most influential; no parameter made zirconia cost-effective. PSA: zirconia cost-effective in 0.50% of iterations at £20,000. Discount rate scenarios: 1% ICER £232,707; 5% ICER £372,014. Copayment at 75%: ICER £41,741. |
| **Effect of engagement with patients and others affected by the study** | 25 | Report on any difference patient/service recipient, general public, community, or stakeholder involvement made to the approach or findings of the study | Not applicable. No formal engagement reported (see Item 21). |
| **Discussion** |  |  |  |
| **Study findings, limitations, generalisability, and current knowledge** | 26 | Report key findings, limitations, ethical or equity considerations not captured, and how these could affect patients, policy, or practice. | Section 4.1–4.5 (pp. 41–45): Key finding: £5,875 premium for zirconia with marginal QALY gain (0.019). Limitations: retrospective evidence only, no RCTs, short follow-up, heterogeneous material reporting, no ISFCDP-specific utility values. Patient choice framed as trade-off between fewer but more consequential zirconia complications vs. more frequent but chairside-repairable acrylic complications. |
| **Other relevant information** |  |  |  |
| **Source of funding** | 27 | Describe how the study was funded and any role of the funder in the identification, design, conduct, and reporting of the analysis | Declarations – Funding (p. 46): No specific funding received. RV (CEO of 21D Clinical, employer of EC and NP) had no role in data collection, analysis, interpretation, or decision to submit. Cost data provided by 21D declared as methodological advantage. |
| **Conflicts of interest** | 28 | Report authors conflicts of interest according to journal or International Committee of Medical Journal Editors requirements. | Declarations – Competing interests (p. 46): EC and NP employed by 21D Clinical Limited, which manufactures CAD/CAM titanium-PMMA ISFCDPs. Systematic review and CEA conducted independently. RV had no role in analysis or interpretation. |

Supplementary Table S4 – Full search strings

| **Block** | **Search terms** |
| --- | --- |
| Dental implants and complete arch | ("dental implants"[MeSH] OR "dental implant*" OR "implant-supported" OR "implant supported" OR "complete arch" OR "complete-arch" OR "full arch" OR "full-arch" OR "All-on-4" OR "All-on-four" OR "All-on-6" OR "All-on-six" OR "edentulous" OR "fixed complete denture" OR "fixed dental prosthesis" OR "ISFC*" OR "hybrid prostheses*") |
| Zirconia / monolithic | ("zirconia" OR "zirconium dioxide" OR "ZrO2" OR "monolithic" OR "Y-TZP" OR "yttria" OR "ceramic prostheses*") |
| Metal framework and PMMA/acrylic | ("titanium" OR "metal framework" OR "metal-acrylic" OR "metal acrylic" OR "PMMA" OR "polymethyl methacrylate" OR "acrylic" OR "acrylic resin" OR "resin teeth" OR "hybrid prostheses*" OR "cobalt-chromium" OR "Co-Cr") |

Supplementary Tables S5 – Full costs breakdown

| **Unit rates** | **Value** | **Source** |
| --- | --- | --- |
| Median hourly wage (April 2025) | £19.67 | ONS ASHE 2025 |
| HMRC mileage rate (£/mile, cars & vans) | £0.45 | HMRC 2025 |
| Average round-trip distance to specialist clinics | 60 | Assumption |

| **UK market survey of full-arch implant-supported All-on-4 prosthesis costs (search made on March 2026)** | | | |
| --- | --- | --- | --- |
| **Source** | **Ti-PMMA** | **Zirconia** | **Notes** |
| A (mint dental clinic) | - | - | Emailed no response |
| B (Evo dental) | £15,500 | £16,500 |  |
| C (21D Clinical) | £16,995 | - |  |
| D (Dental by design) | - | £20,990 |  |
| E (Yorkshire dental suite) | £19,995 | £29,995 |  |
| F (Smiledent Dental & Implant Centre) |  | £30,000 |  |
| G (Zental) | - | £23,980 |  |

Consistent with NICE methodology when no published list price or national price is agreed.

| **Direct healthcare costs** | **Ti-PMMA** | **Zirconia** | **Source** |
| --- | --- | --- | --- |
| Initial placement surgery | £17,497 | £24,293 | UK average of survey |
| Annual maintenance visit | £250 | £250 | UK survey |
| Minor repair (chairside / short lab work) | £350 | £500 |  |
| Major remake (full framework and teeth fabrication) | £2,500 | £5,000 |  |
| Implant replacement (surgical and prosthetic refit) | £2,500 | £2,500 |  |
| Prosthesis failure (ongoing annual denture associated cost est.) | £500 | £500 |  |

| **Patient time off work (hours per event)** | **Ti-PMMA** | **Zirconia** | **Note** |
| --- | --- | --- | --- |
| Initial placement surgery | 16 | 16 |  |
| Annual maintenance visit | 3 | 3 |  |
| Minor repair (chairside / short lab work) | 3 | 3 |  |
| Major remake (full framework and teeth fabrication) | 16 | 16 |  |
| Implant replacement (surgical and prosthetic refit) | 16 | 16 |  |
| Prosthesis failure (ongoing annual denture associated cost est.) | 2 | 2 |  |

| **Travel cost per event (miles * mileage rate)** | **Ti-PMMA** | **Zirconia** |
| --- | --- | --- |
| Initial placement surgery | £27.00 | £27.00 |
| Annual maintenance visit | £27.00 | £27.00 |
| Minor repair (chairside / short lab work) | £27.00 | £27.00 |
| Major remake (full framework and teeth fabrication) | £108.00 | £108.00 |
| Implant replacement (surgical and prosthetic refit) | £81.00 | £81.00 |
| Prosthesis failure (annual) | £27.00 | £27.00 |

| **Productivity loss per event (hours * median wage)** | **Ti-PMMA** | **Zirconia** |
| --- | --- | --- |
| Initial placement surgery | £314.72 | £314.72 |
| Annual maintenance visit | £59.01 | £59.01 |
| Minor repair (chairside / short lab work) | £59.01 | £59.01 |
| Major remake (full framework and teeth fabrication) | £314.72 | £314.72 |
| Implant replacement (surgical and prosthetic refit) | £314.72 | £314.72 |
| Prosthesis failure (annual) | £39.34 | £39.34 |

| **TOTAL PATIENT COST PER EVENT** | **Ti-PMMA** | **Zirconia** | **Components** |
| --- | --- | --- | --- |
| Initial placement surgery | £17,838.39 | £24,634.72 | Direct + productivity loss + travel |
| Annual maintenance visit | £336.01 | £336.01 |  |
| Minor repair (chairside / short lab work) | £436.01 | £586.01 |  |
| Major remake (full framework and teeth fabrication) | £2,922.72 | £5,422.72 |  |
| Implant replacement (surgical and prosthetic refit) | £2,895.72 | £2,895.72 |  |
| Prosthesis failure (ongoing annual denture associated cost est.) | £566.34 | £566.34 |  |

Supplementary Table S6 – Full utility value justification

| **Disutility Parameter** | **Base case** | **Range (SA)** | **Distribution** | **Duration** | **Literature evidence** |
| --- | --- | --- | --- | --- | --- |
| Minor complication (state 2) | -0.05 | -0.01 to -0.10 | Beta | 1-4 weeks  (0.038 years) | There is no ISFCDP-specific ED-5D data so values had to be estimated [1]. A NICE wisdom tooth HTA used a disutility of -0.345 from Ara & Brazier’s estimations for active symptomatic disease with pain [2, 3]. A minor prosthetic complication will involve brief discomfort compared to active dental disease. We therefore applied one-seventh of the Ara & Brazier oral-condition decrement, which is consistent with the NICE wisdom tooth model’s approach pf halving the full decrement. This is varied widely in sensitivity analysis. |
| Major complication (state 3) | -0.15 | -0.10 to -0.25 | Beta | 4-12 weeks (0.13 years) | While a prosthesis is undergoing a remake, the patient will be edentulous or wearing a removal temporary prosthesis. A Brazilian cost-effectiveness study used a value of 0.94 for ISFCDPs (against a value of 0.79 for conventional dentures) although this was to build quality-adjusted prosthetic years (QAPY) [4}. These values are used in a Markov model over 20 years. We then used this 16% relative difference to choose a 10% multiplicative decrement as a conservative estimate as QAPY captures oral-specific utility while EQ-5D has a lower sensitivity to oral states. |
| Implant loss (state 4) | -0.20 | -0.10 to -0.30 | Beta | 1.5-6 months  (0.25 years) | Surgical reintervention, an additional healing period and prosthodontic management are needed in this state. The NICE wisdom tooth HTA used the full -0.345 oral condition decrement for extraction complications. We decided on 60% of this value reflecting the need for surgical reintervention and recovery, but without the pain of impacted third molar complications. A Britain-based study found that edentulous patients with inadequate prosthesis were 2.66 times more likely to report oral impacts , supporting the use of a strong time-limited disutility value [5]. |
| Prosthesis failure multiplier (state 5) | 0.10 | 0.05 to 0.15 | Beta | Permanent | We determined a 10% multiplier as relatively conservative to other disutility decrements we have used. This is because we recognise that QAPY are not EQ-5D based and we have to consider QALYs low sensitivity to oral impacts. |

Table S7 – Justification for CEA transition probabilities

| Transition | Ti-PMMA value | Zirconia value | Ti-PMMA reference | Zirconia reference | Justification |
| --- | --- | --- | --- | --- | --- |
| S1→S2: Functioning→Minor complication | 12.0% | 3.0% | Barootchi, 2020 | [6]; [7] | Barootchi metal-acrylic reported 94 tooth fracture/chipping events over 8.7 years = 0.251, but included repeat events on the same prosthesis, so adjusted for that. Zirconia arm reported near zero and infrequent chipping events in purely monolithic zirconia prosthetics. Capparé and Durrani saw zero events in 3 and 2 years of follow-up |
| S1→S3: Functioning→Major complication | 4.0% | 0.3% | Barootchi, 2020 | [6]; [7] | Metal-acrylic reported 83% prosthesis survival at 5 years = r = -ln(0.83)/5 = 0.037, rounded to 0.04. Lower survival was recorded but this was in a teaching institution. Zirconia arm was derived from two fracture rates; 2 framework fractures in 115 prostheses over 5.2 years = 0.003, and 6 fractures in 2,039 prostheses over 5 years = 0.001. |
| S1→S4: Functioning→Implant loss | 0.5% | 0.5% | Barootchi, 2020; [8] | Barootchi, 2020; [9] | No statistical difference in implant failure between zirconia and metal-acrylic arms. Implant survival at 5 years and over is reported to be +95%. |
| S1→S5: Functioning→Prosthesis failure | 0.2% | 0.2% | Assumption | Assumption | Nearly all prosthesis failure events in reality will be due to other complications, minor or major, first. |
| S2→S1: Minor comp→Functioning | 100.0% | 100.0% | Assumption | Assumption | Due to our definition of minor complications, these are fixed by chairside repair or short laboratory intervention. |
| S3→S1: Major comp→Functioning | 95.0% | 95.0% | Barootchi, 2020 | Barootchi, 2020 | The majority of patients receiving a remake will return to full function. Set equal for both arms as a successful remake is determined by clinical and patient factors, not material type. |
| S3→S5: Major comp→Failure | 5.0% | 5.0% | Barootchi, 2020 | Barootchi, 2020 | From the results that 5% of prostheses experiencing major complications were not remade. |
| S4→S1: Implant loss→Functioning | 70.0% | 70.0% | Assumption | Assumption | Majority of single implant loss events are recoverable, except in events where bone position is lost or bone volume is insufficient for replacement. |
| S4→S5: Implant loss→Failure | 10.0% | 10.0% | [10] | [10] | Mackert et al. report that 7% of total prosthesis replacements were due to implant loss, but this was for cases where loss was the primary cause. 10% was chosen as a conservative estimate. |
| S5→S5: Failure→Failure (absorbing) | 100.0% | 100.0% | Assumption | Assumption | State modelled as an absorbing state. Patients either transition to removeable dentures or edentulism. Simplifying assumption. |

Table S8 – Studies excluded from GRADE Evidence Profile and reason

This does not include studies that are automatically excluded due to not reporting the outcome.

| **Outcome** | **Study excluded** | **Reason for exclusion** |
| --- | --- | --- |
| Prosthesis survival | Box et al., 2021 | Complication-free prostheses reported rather than true, consistent prosthesis survival/failure |
|  | Sabău et al., 2023 | Mixed prosthetic design with no direct zirconia vs metal-framework comparison, combines fixed and removable prostheses |
|  | Durrani et al., 2020 | Survival outcomes not reported by prosthesis material |
| Technical complications (overall) | Sabău et al., 2023 | Technical complication outcomes reported across fixed and removeable prosthesis types |
|  | Durrani et al., 2020 | Qualitatively reported without clear, extractable comparative counts |
| Framework fracture | Sabău et al., 2023 | Complications not split by zirconia versus comparator |
|  | Durrani et al., 2020 | Framework fracture outcomes not distinctly reported |
| Veneer fracture/chipping | Sabău et al., 2023 | Not separated by prosthesis material group |
|  | Durrani et al., 2020 | Not in an extractable format |
| Biological complications | Capparé et al., 2021 | Study used peri-implant soft-tissue indices rather than discrete complication events |
|  | Anya et al., 2021 | Defined biological complications using peri-implant indices |
| Implant survival | Sabău et al., 2023 | Not stratified by prosthesis material |
|  | Box et al., 2021 | Not reported by prosthesis group in a comparable format |
| Plaque index | Tang et al., 2019 | Not reported as comparable quantitative outcomes |
| Bleeding on probing | Tang et al., 2019 | Not reported as comparable quantitative outcomes |
| Probing depth | Tang et al., 2019 | Not reported as comparable quantitative outcomes |
| Patient satisfaction / OHRQoL | Tang et al., 2019 | Not reported as comparable quantitative outcomes |
| Patient satisfaction / OHRQoL | Capparé et al., 2021 | Not reported |

Table S9 – Net monetary benefit for zirconia

NMB at £20,000/QALY

|  |  |
| --- | --- |
| 10000 | -5688.43 |
| 20000 | -5502.18 |
| 30000 | -5315.93 |
| 40000 | -5129.68 |
| 50000 | -4943.42 |
| 60000 | -4757.17 |
| 70000 | -4570.92 |
| 80000 | -4384.67 |
| 90000 | -4198.42 |
| 100000 | -4012.17 |
| 110000 | -3825.92 |
| 120000 | -3639.67 |
| 130000 | -3453.41 |
| 140000 | -3267.16 |
| 150000 | -3080.91 |
